# Collateral effects of the COVID-19 pandemic on endocrine treatments for breast and prostate cancer in the UK: implications for bone health

**DOI:** 10.1101/2023.11.09.23298305

**Authors:** Nicola L Barclay, Marti Català, Annika M. Jödicke, Daniel Prieto-Alhambra, Danielle Newby, Antonella Delmestri, Wai Yi Man, Àlvar Roselló Serrano, Marta Pineda Moncusí, The OPTIMA Consortium

## Abstract

**Background:** The COVID-19 pandemic affected cancer screening, diagnosis and treatment pathways. This study examined the impact of the pandemic on incidence and trends of endocrine treatments in patients with breast or prostate cancer; and endocrine treatment-related side-effects.

**Methods:** Population-based cohort study using UK primary care Clinical Practice Research Datalink (CPRD) GOLD database. There were 13,701 newly diagnosed breast cancer patients and 12,221 prostate cancer patients with ≥1-year data availability since diagnosis between January 2017-June 2022. Incidence rates (IR) and incidence rate ratios (IRR) were calculated across multiple time periods before and after lockdown to examine the impact of changing social restrictions on endocrine treatments and treatment-related outcomes, including osteopenia, osteoporosis and bisphosphonate prescriptions.

**Results:** In patients with breast cancer, aromatase inhibitor prescriptions increased during lockdown compared to pre-pandemic (IRR: 1.22 [95% Confidence Interval: 1.11-1.34]), followed by a decrease post-first lockdown (IRR: 0.79 [95%CI: 0.69-0.89]). In patients with prostate cancer, first-generation antiandrogen prescriptions increased compared to pre-pandemic (IRR: 1.23 [95% CI: 1.08-1.4]). For breast cancer patients on aromatase inhibitors, diagnoses of osteopenia, osteoporosis and bisphosphonate prescriptions were reduced across all lockdown periods compared to pre-pandemic (IRR range: 0.31-0.62).

**Conclusion:** During the first two years of the pandemic, newly diagnosed breast and prostate cancer patients were prescribed more endocrine treatments compared to pre-pandemic, due to restrictions on hospital procedures replacing surgeries with bridging therapies. But breast cancer patients had fewer diagnoses of osteopenia and osteoporosis, and bisphosphonate prescriptions. These patients should be followed up in the coming years for signs of bone thinning. Evidence of poorer management of treatment-related side-effects will allow us to determine whether there is a need to better allocate resources to patients at high risk for bone-related complications.

## Background

The COVID-19 pandemic affected healthcare beyond the immediate effects of the virus. The collateral impact of lockdown affected cancer screening, diagnosis and treatment pathways, ultimately decreasing cancer survival [1]. Indeed, recent reports highlight that screening tests for breast cancer and visits to breast surgeons were delayed in the initial months following lockdown and up to at least June 2022 in the United Kingdom (UK) [2, 3]. Furthermore, breast and prostate cancer were under-diagnosed between March 2020 and June 2022 [2, 4].

Because healthcare staff were redeployed to care for COVID-19 patients, and many hospital beds were allocated to such patients, treatments for cancers were altered [4]. New guidelines were introduced in Europe for the management of cancer patients during the pandemic, including the recommendation to postpone surgery/radiotherapy and instead provide neo-adjuvant endocrine therapy for some breast cancer patients during the waiting period (though not specifically in the UK) [5]. Similar recommendations were implemented for prostate cancer. Patients with intermediate or high-risk prostate cancer were recommended to delay radiotherapy or surgical treatment for 3-6 months and instead be administered androgen deprivation therapy (ADT) during this waiting period in some European countries [5].

Increases in the use of endocrine therapies during the initial phases of the pandemic enabled access to treatment amidst a period of turmoil. Nevertheless, consideration of the side-effects of these treatments, and how such side-effects were managed during the pandemic, cannot be neglected. Indeed, well-known side-effects of endocrine therapies such as aromatase inhibitors (AIs) for breast cancer, gonadotropin-releasing hormone (GnRH) analogues for breast and prostate cancer, and ADT for prostate cancer, include musculoskeletal problems such as bone density loss, osteopenia, and osteoporosis, increasing the risk of bone fractures in patients exposed to such drugs [6, 7]. Common preventative and treatment strategies to improve bone health in such patients include the administration of anti-osteoporotic treatments. However, the assessment of endocrine therapy side-effects such as osteopenia and osteoporosis were conceivably not a priority during the COVID-19 pandemic. Thus, subsequent diagnosis and treatment of treatment-related conditions due to these therapies may have decreased during the COVID-19 pandemic. Despite this hypothesis, there is no available data on the pandemic’s impact on secondary diagnoses and anti-osteoporotic treatment prescriptions in breast and prostate cancer patients.

The primary aim of this study is to examine how the changing social restrictions imposed by the pandemic affected incidence and trends of endocrine treatment prescriptions in newly diagnosed (incident) breast and prostate cancer patients; and secondarily, endocrine treatment-related outcomes (including prescriptions of bisphosphonates, osteopenia, and osteoporosis), in UK clinical practice from March 2020 to June 2022. Evidence of poorer management of treatment-related side-effects will allow us to determine whether there is a need to better allocate resources to patients at high risk for bone-related complications.

## Methods

### Study design and participants

This is a population-based cohort study using routinely collected electronic health records from UK Clinical Practice Research Datalink (CPRD) GOLD. CPRD GOLD contains pseudo-anonymised patient-level demographics, lifestyle data, clinical diagnoses, prescriptions and preventive care contributed by general practitioners (GP) from the UK [8]. The use of CPRD data for this study was approved by the Independent Scientific Advisory Committee (22_002331). This database was mapped to the Observational Medical Outcomes Partnership (OMOP) Common Data Model (CDM) [9].

People were eligible if they were registered between January 2017 and June 2022 with at least one year of data availability in the database before their cancer diagnosis. The incident breast and prostate cancer cohorts excluded individuals diagnosed with the same cancer any time in clinical history; and those with metastases, as we were interested in the pandemic’s effect on cancer patients who had not previously been under cancer management pathways. All endocrine treatments and treatment-related outcomes were first-ever events in clinical history.

### Drug Utilisation

The study focused on prescriptions of AIs, AIs with GnRH agonists or antagonists, Tamoxifen, and Tamoxifen with GnRH agonists or antagonists in breast cancer patients; and first-generation ADT, GnRH agonists, GnRH agonists with first generation ADT, GnRH antagonists, and second-generation ADT in prostate cancer patients. Endocrine treatment-related side-effects in breast and prostate cancer patients included prescriptions of bisphosphonates, osteopenia, and osteoporosis.

All cancer diagnoses and medications were defined based on Systematized Nomenclature of Medicine Clinical Terms (SNOMED CT) / RxNorm codes (as appropriate), in the OMOP-mapped data. Diagnostic codes indicative of either non-malignant cancer or metastasis were. The cancer diagnosis definitions and endocrine treatments were reviewed with the aid of the CohortDiagnostics R package [10]. This package was used to identify additional codes of interest and to remove those highlighted as irrelevant based on feedback from clinicians with oncology expertise through an iterative process during the initial stages of analysis. A list of all codes used to define the population and each outcome can be found in our Github repository: https://github.com/oxford-pharmacoepi/CancerCovidEndocrineTx/tree/main/Concept%20Sets

### Public Health Restrictions

The ‘exposures’ were the periods of the changing social restrictions due to the pandemic in the UK. Our observation period was dissected into seven time-periods as follows: pre-pandemic (January 2017 to February 2020), first lockdown (March 2020 to June 2020), post-first lockdown (July 2020 to October 2020), second lockdown (November 2020 to December 2020), third lockdown (January 2021 to March 2021), easing of restrictions (April 2021 to June 2021), and legal restrictions removed (July 2021 to June 2022). We also examined the period covering all lockdown periods from March 2020 to June 2022 to make comparisons with the pre-pandemic period.

## Statistical analyses

### Characterisation

Patients with incident endocrine treatment prescriptions were characterised on age at index date (date of incident outcome), sex, comorbidities (based on SNOMED codes) any time in patient history, and medication use (based on RxNorm codes) within the 90 days prior to their first endocrine prescription to gain an understanding of their clinical profile. Continuous variables were summarised as means and standard deviations, and medians and interquartile ranges, and categorical variables as counts and percentages. Frequency counts less than five were censored to enhance patient/practice confidentiality.

### Incidence Rates and Incidence Rate Ratios

Incidence rates (IR) with 95% confidence intervals (CI) were calculated for all endocrine treatments and treatment-related outcomes monthly, and within the pre-pandemic, lockdown, and post-lockdown periods across the entire observation period using the IncidencePrevalence R package [11]. Patients with breast cancer or prostate cancer who were diagnosed within the observation period contributed time-at-risk, and as such contributed to the ‘denominator population’, until the earliest of a record of the endocrine treatment / treatment-related outcome, transfer out of the database, end of the study period or death. Incidence rate ratios (IRR) with 95% CI were calculated using the IR estimates across the post-lockdown periods divided by the IR estimates before lockdown. All statistical code can be found in our Github repository: https://github.com/oxford-pharmacoepi/CancerCovidEndocrineTx

## Results

### Characterisations of breast and prostate cancer patients

Overall, there were 13,760 incident breast cancer patients, and 8,805 incident prostate cancer patients included in the denominator populations from January 2017 to June 2022. Of those, there were 8,805 breast cancer patients, and 8,591 prostate cancer patients on endocrine treatments in the year after diagnosis. These patients may have been prescribed more than one endocrine treatment during this period after diagnosis. Attrition tables showing how the study cohorts were derived are shown in **Supplementary Tables S1 and S2**.

Demographic characteristics, comorbidities and comedications in the breast and prostate cancer patients on different endocrine treatments are shown in **Table 1** and **Table 2**. Breast cancer patients on AIs were older and had a greater proportion of comorbidities and comedications compared to the other breast cancer patient groups. There were no patterns in the comorbidities or comedications of prostate cancer patients as a function of their endocrine treatment.

**Table 1.** Characterisations of breast cancer patients on endocrine treatments.

|  | **Aromatase Inhibitors** | **Aromatase Inhibitors**<br>**with GnRH Agonists or**<br>**Antagonists** | **Tamoxifen** | **Tamoxifen with GnRH**<br>**Agonists or Antagonists** |
| --- | --- | --- | --- | --- |
| N records | 6634 | 253 | 2887 | 132 |
| Mean (SD) age | 68.2 (11.97) | 44.86 (6.27) | 54.69 (12.19) | 42.94 (6.94) |
| Median (IQR) age | 68 (59-77) | 46 (40-50) | 52 (47-62) | 44 (38.75-48) |
| Median prior history in years (IQR) | 16.06 (11.1-18.85) | 14.47 (6.65-18.23) | 15.15 (9.48-17.95) | 13.86 (6.66-17.46) |
| N male | 0 | 0 | 0 | 0 |
| N Female | 6634 | 253 | 2887 | 132 |
| % Female | 100% | 100% | 100% | 100% |
| **Comorbidities (n(%))** |  |  |  |  |
| Atrial Fibrillation | 390 (5.88%) | 0 (0%) | 40 (1.39%) | 0 (0%) |
| Cerebrovascular Disease | 306 (4.61%) | <5 (~1%) | 35 (1.21%) | <5 (~1%) |
| Chronic Liver Disease | 40 (0.6%) | <5 (~0%) | 8 (0.28%) | 0 (0%) |
| Chronic Obstructive Lung Disease | 348 (5.25%) | <5 (~0%) | 80 (2.77%) | 0 (0%) |
| Coronary Arteriosclerosis | 38 (0.57%) | 0 (0%) | 13 (0.45%) | 0 (0%) |
| Crohn's Disease | 18 (0.27%) | 0 (0%) | 8 (0.28%) | 0 (0%) |
| Dementia | 127 (1.91%) | 0 (0%) | 10 (0.35%) | 0 (0%) |
| Depressive Disorder | 1195 (18.01%) | 58 (22.92%) | 540 (18.7%) | 29 (21.97%) |
| Diabetes Mellitus | 731 (11.02%) | 11 (4.35%) | 127 (4.4%) | <5 (~3%) |
| H. Pylori Infection | 20 (0.3%) | <5 (~0%) | <5 (~0%) | 0 (0%) |
| Heart Disease | 925 (13.94%) | 5 (1.98%) | 172 (5.96%) | <5 (~1%) |
| Heart Failure | 129 (1.94%) | 0 (0%) | 14 (0.48%) | 0 (0%) |
| Hepatitis C | <5 (~0%) | 0 (0%) | <5 (~1%) | 0 (0%) |
| HIV | 0 (0%) | 0 (0%) | 0 (0%) | 0 (0%) |
| Hyperlipidemia | 615 (9.27%) | <5 (~1%) | 109 (3.78%) | <5 (~1%) |
| Hypertension | 1835 (27.66%) | 26 (10.28%) | 442 (15.31%) | 10 (7.58%) |
| Ischemic Heart Disease | 324 (4.88%) | <5 (~1%) | 62 (2.15%) | 0 (0%) |
| Lesion Liver | 87 (1.31%) | 9 (3.56%) | 16 (0.55%) | 0 (0%) |
| Obesity | 277 (4.18%) | 6 (2.37%) | 78 (2.7%) | <5 (~3%) |
| Osteoarthritis | 1532 (23.09%) | <5 (~1%) | 328 (11.36%) | <5 (~2%) |
| Peripheral Vascular Disease | 81 (1.22%) | 0 (0%) | 7 (0.24%) | 0 (0%) |
| Pneumonia | 142 (2.14%) | <5 (~1%) | 36 (1.25%) | 0 (0%) |

|  | **Aromatase Inhibitors** | **Aromatase Inhibitors**<br>**with GnRH Agonists or**<br>**Antagonists** | **Tamoxifen** | **Tamoxifen with GnRH**<br>**Agonists or Antagonists** |
| --- | --- | --- | --- | --- |
| Psoriasis | 245 (3.69%) | 11 (4.35%) | 98 (3.39%) | 5 (3.79%) |
| Pulmonary Embolism | 119 (1.79%) | <5 (~1%) | 12 (0.42%) | 0 (0%) |
| Renal Impairment | 811 (12.22%) | <5 (~1%) | 124 (4.3%) | 0 (0%) |
| Rheumatoid Arthritis | 66 (0.99%) | <5 (~0%) | 30 (1.04%) | <5 (~1%) |
| Schizophrenia | 21 (0.32%) | <5 (~0%) | 5 (0.17%) | 0 (0%) |
| Ulcerative Colitis | 29 (0.44%) | 0 (0%) | 9 (0.31%) | <5 (~1%) |
| UTI Disease | 1055 (15.9%) | 33 (13.04%) | 375 (12.99%) | 10 (7.58%) |
| Venous Thrombosis | 418 (6.3%) | 15 (5.93%) | 93 (3.22%) | <5 (~3%) |
| Visual System Disorder | 2437 (36.74%) | 48 (18.97%) | 731 (25.32%) | 20 (15.15%) |
| **Comedications (n(%))** |  |  |  |  |
| Antidepressants | 3529 (53.2%) | 146 (57.71%) | 1468 (50.85%) | 58 (43.94%) |
| Antiepileptics | 1214 (18.3%) | 32 (12.65%) | 451 (15.62%) | 17 (12.88%) |
| Antiinflammatory / Antirheumatic | 4489 (67.67%) | 155 (61.26%) | 1930 (66.85%) | 76 (57.58%) |
| Antineoplastics | 426 (6.42%) | <5 (~1%) | 122 (4.23%) | 5 (3.79%) |
| Antipsoriatics | 147 (2.22%) | 9 (3.56%) | 77 (2.67%) | <5 (~3%) |
| Antipsychotics | 2020 (30.45%) | 77 (30.43%) | 785 (27.19%) | 37 (28.03%) |
| Antithrombotics | 1256 (18.93%) | 32 (12.65%) | 218 (7.55%) | 13 (9.85%) |
| Anxiolytics | 2115 (31.88%) | 86 (33.99%) | 888 (30.76%) | 40 (30.3%) |
| Beta Blockers | 2055 (30.98%) | 68 (26.88%) | 725 (25.11%) | 34 (25.76%) |
| Calcium Channel Blockers | 2092 (31.53%) | 18 (7.11%) | 421 (14.58%) | 7 (5.3%) |
| Diuretics | 2267 (34.17%) | 18 (7.11%) | 449 (15.55%) | 6 (4.55%) |
| Drugs For Acid Related Disorders | 4496 (67.77%) | 141 (55.73%) | 1707 (59.13%) | 67 (50.76%) |
| Drugs For Diabetes | 660 (9.95%) | 13 (5.14%) | 134 (4.64%) | <5 (~3%) |
| Drugs For Obstructive Airway<br>Diseases | 3239 (48.82%) | 126 (49.8%) | 1350 (46.76%) | 60 (45.45%) |
| Hypnotics / Sedatives | 1686 (25.41%) | 77 (30.43%) | 778 (26.95%) | 39 (29.55%) |
| Immunosuppressants | 128 (1.93%) | <5 (~1%) | 51 (1.77%) | <5 (~1%) |
| Opioids | 4517 (68.09%) | 160 (63.24%) | 1798 (62.28%) | 79 (59.85%) |
| Psychostimulants | <5 (~1%) | 0 (0%) | <5 (~0%) | 0 (0%) |

Note. IQR = Interquartile range; SD = standard deviation; counts <5 masked and proportions rounded to nearest 1% in order patients to remain masked.

**Table 2.**
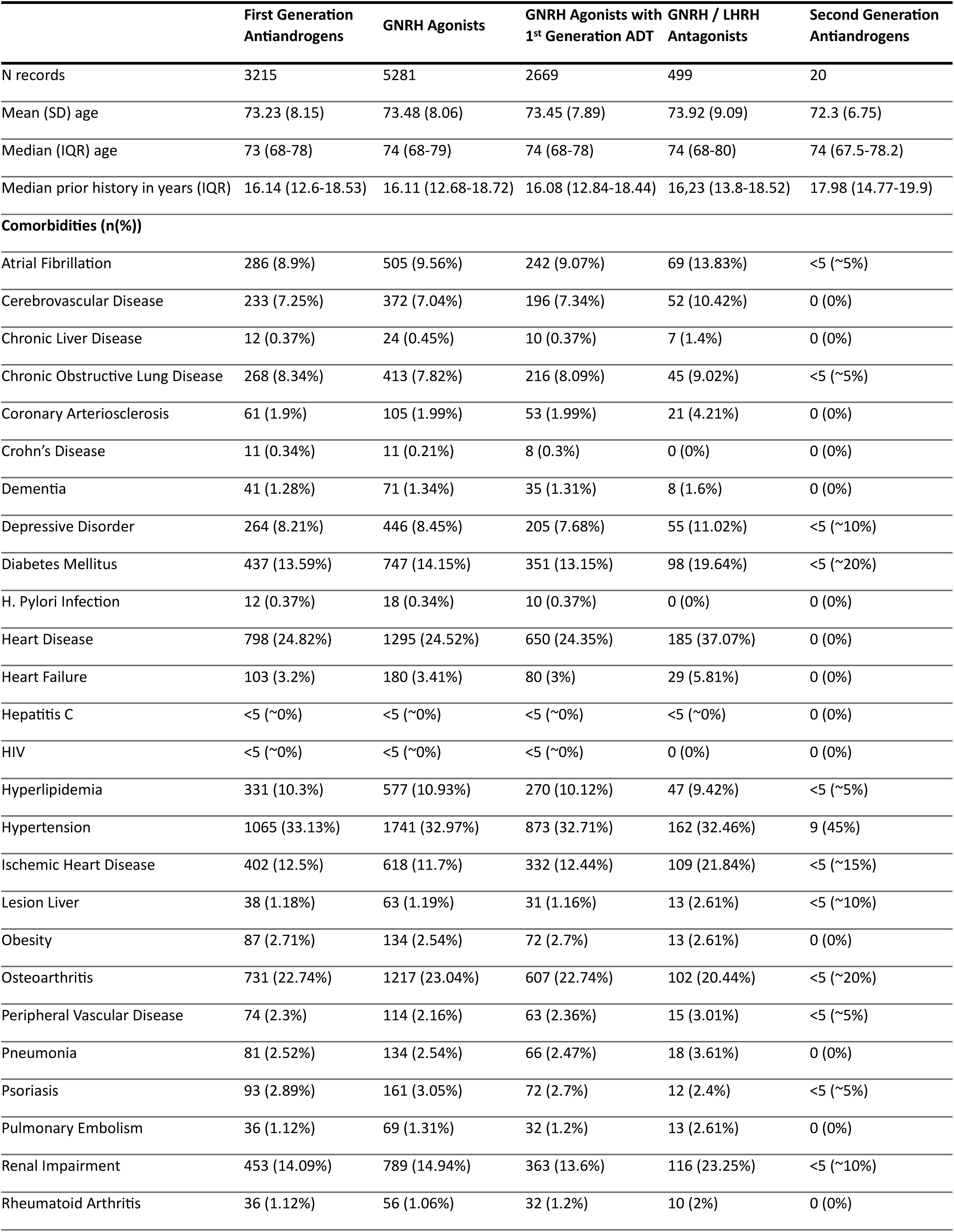
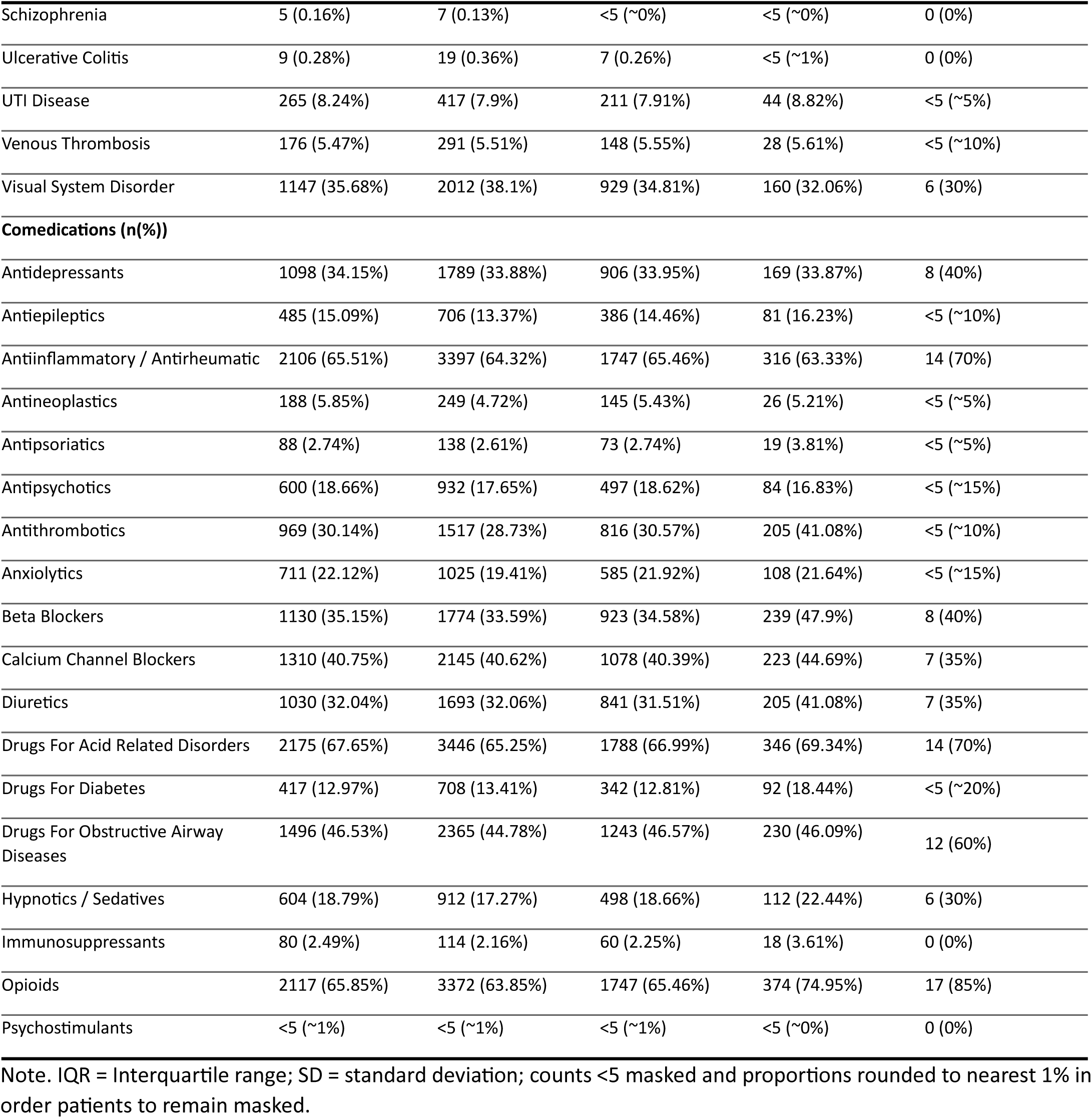
Characterisations of prostate cancer patients on endocrine treatments.

|  | First Generation<br>Antiandrogens | GNRH Agonists | GNRH Agonists with<br>1<sup>st</sup> Generation ADT | GNRH / LHRH<br>Antagonists | Second Generation<br>Antiandrogens |
| --- | --- | --- | --- | --- | --- |
| N records | 3215 | 5281 | 2669 | 499 | 20 |
| Mean (SD) age | 73.23 (8.15) | 73.48 (8.06) | 73.45 (7.89) | 73.92 (9.09) | 72.3 (6.75) |
| Median (IQR) age | 73 (68-78) | 74 (68-79) | 74 (68-78) | 74 (68-80) | 74 (67.5-78.2) |
| Median prior history in years (IQR) | 16.14 (12.6-18.53) | 16.11 (12.68-18.72) | 16.08 (12.84-18.44) | 16,23 (13.8-18.52) | 17.98 (14.77-19.9) |
| **Comorbidities (n(%))** |  |  |  |  |  |
| Atrial Fibrillation | 286 (8.9%) | 505 (9.56%) | 242 (9.07%) | 69 (13.83%) | <5 (~5%) |
| Cerebrovascular Disease | 233 (7.25%) | 372 (7.04%) | 196 (7.34%) | 52 (10.42%) | 0 (0%) |
| Chronic Liver Disease | 12 (0.37%) | 24 (0.45%) | 10 (0.37%) | 7 (1.4%) | 0 (0%) |
| Chronic Obstructive Lung Disease | 268 (8.34%) | 413 (7.82%) | 216 (8.09%) | 45 (9.02%) | <5 (~5%) |
| Coronary Arteriosclerosis | 61 (1.9%) | 105 (1.99%) | 53 (1.99%) | 21 (4.21%) | 0 (0%) |
| Crohn's Disease | 11 (0.34%) | 11 (0.21%) | 8 (0.3%) | 0 (0%) | 0 (0%) |
| Dementia | 41 (1.28%) | 71 (1.34%) | 35 (1.31%) | 8 (1.6%) | 0 (0%) |
| Depressive Disorder | 264 (8.21%) | 446 (8.45%) | 205 (7.68%) | 55 (11.02%) | <5 (~10%) |
| Diabetes Mellitus | 437 (13.59%) | 747 (14.15%) | 351 (13.15%) | 98 (19.64%) | <5 (~20%) |
| H. Pylori Infection | 12 (0.37%) | 18 (0.34%) | 10 (0.37%) | 0 (0%) | 0 (0%) |
| Heart Disease | 798 (24.82%) | 1295 (24.52%) | 650 (24.35%) | 185 (37.07%) | 0 (0%) |
| Heart Failure | 103 (3.2%) | 180 (3.41%) | 80 (3%) | 29 (5.81%) | 0 (0%) |
| Hepatitis C | <5 (~0%) | <5 (~0%) | <5 (~0%) | <5 (~0%) | 0 (0%) |
| HIV | <5 (~0%) | <5 (~0%) | <5 (~0%) | 0 (0%) | 0 (0%) |
| Hyperlipidemia | 331 (10.3%) | 577 (10.93%) | 270 (10.12%) | 47 (9.42%) | <5 (~5%) |
| Hypertension | 1065 (33.13%) | 1741 (32.97%) | 873 (32.71%) | 162 (32.46%) | 9 (45%) |
| Ischemic Heart Disease | 402 (12.5%) | 618 (11.7%) | 332 (12.44%) | 109 (21.84%) | <5 (~15%) |
| Lesion Liver | 38 (1.18%) | 63 (1.19%) | 31 (1.16%) | 13 (2.61%) | <5 (~10%) |
| Obesity | 87 (2.71%) | 134 (2.54%) | 72 (2.7%) | 13 (2.61%) | 0 (0%) |
| Osteoarthritis | 731 (22.74%) | 1217 (23.04%) | 607 (22.74%) | 102 (20.44%) | <5 (~20%) |
| Peripheral Vascular Disease | 74 (2.3%) | 114 (2.16%) | 63 (2.36%) | 15 (3.01%) | <5 (~5%) |
| Pneumonia | 81 (2.52%) | 134 (2.54%) | 66 (2.47%) | 18 (3.61%) | 0 (0%) |
| Psoriasis | 93 (2.89%) | 161 (3.05%) | 72 (2.7%) | 12 (2.4%) | <5 (~5%) |
| Pulmonary Embolism | 36 (1.12%) | 69 (1.31%) | 32 (1.2%) | 13 (2.61%) | 0 (0%) |
| Renal Impairment | 453 (14.09%) | 789 (14.94%) | 363 (13.6%) | 116 (23.25%) | <5 (~10%) |
| Rheumatoid Arthritis | 36 (1.12%) | 56 (1.06%) | 32 (1.2%) | 10 (2%) | 0 (0%) |

|  | First Generation<br>Antiandrogens | GNRH Agonists | GNRH Agonists with<br>1<sup>st</sup> Generation ADT | GNRH / LHRH<br>Antagonists | Second Generation<br>Antiandrogens |
| --- | --- | --- | --- | --- | --- |
| Schizophrenia | 5 (0.16%) | 7 (0.13%) | <5 (~0%) | <5 (~0%) | 0 (0%) |
| Ulcerative Colitis | 9 (0.28%) | 19 (0.36%) | 7 (0.26%) | <5 (~1%) | 0 (0%) |
| UTI Disease | 265 (8.24%) | 417 (7.9%) | 211 (7.91%) | 44 (8.82%) | <5 (~5%) |
| Venous Thrombosis | 176 (5.47%) | 291 (5.51%) | 148 (5.55%) | 28 (5.61%) | <5 (~10%) |
| Visual System Disorder | 1147 (35.68%) | 2012 (38.1%) | 929 (34.81%) | 160 (32.06%) | 6 (30%) |
| **Comedications (n(%))** |  |  |  |  |  |
| Antidepressants | 1098 (34.15%) | 1789 (33.88%) | 906 (33.95%) | 169 (33.87%) | 8 (40%) |
| Antiepileptics | 485 (15.09%) | 706 (13.37%) | 386 (14.46%) | 81 (16.23%) | <5 (~10%) |
| Antiinflammatory / Antirheumatic | 2106 (65.51%) | 3397 (64.32%) | 1747 (65.46%) | 316 (63.33%) | 14 (70%) |
| Antineoplastics | 188 (5.85%) | 249 (4.72%) | 145 (5.43%) | 26 (5.21%) | <5 (~5%) |
| Antipsoriatics | 88 (2.74%) | 138 (2.61%) | 73 (2.74%) | 19 (3.81%) | <5 (~5%) |
| Antipsychotics | 600 (18.66%) | 932 (17.65%) | 497 (18.62%) | 84 (16.83%) | <5 (~15%) |
| Antithrombotics | 969 (30.14%) | 1517 (28.73%) | 816 (30.57%) | 205 (41.08%) | <5 (~10%) |
| Anxiolytics | 711 (22.12%) | 1025 (19.41%) | 585 (21.92%) | 108 (21.64%) | <5 (~15%) |
| Beta Blockers | 1130 (35.15%) | 1774 (33.59%) | 923 (34.58%) | 239 (47.9%) | 8 (40%) |
| Calcium Channel Blockers | 1310 (40.75%) | 2145 (40.62%) | 1078 (40.39%) | 223 (44.69%) | 7 (35%) |
| Diuretics | 1030 (32.04%) | 1693 (32.06%) | 841 (31.51%) | 205 (41.08%) | 7 (35%) |
| Drugs For Acid Related Disorders | 2175 (67.65%) | 3446 (65.25%) | 1788 (66.99%) | 346 (69.34%) | 14 (70%) |
| Drugs For Diabetes | 417 (12.97%) | 708 (13.41%) | 342 (12.81%) | 92 (18.44%) | <5 (~20%) |
| Drugs For Obstructive Airway<br>Diseases | 1496 (46.53%) | 2365 (44.78%) | 1243 (46.57%) | 230 (46.09%) | 12 (60%) |
| Hypnotics / Sedatives | 604 (18.79%) | 912 (17.27%) | 498 (18.66%) | 112 (22.44%) | 6 (30%) |
| Immunosuppressants | 80 (2.49%) | 114 (2.16%) | 60 (2.25%) | 18 (3.61%) | 0 (0%) |
| Opioids | 2117 (65.85%) | 3372 (63.85%) | 1747 (65.46%) | 374 (74.95%) | 17 (85%) |
| Psychostimulants | <5 (~1%) | <5 (~1%) | <5 (~1%) | <5 (~0%) | 0 (0%) |

Note. IQR = Interquartile range; SD = standard deviation; counts <5 masked and proportions rounded to nearest 1% in order patients to remain masked.

### Incidence rates of endocrine treatments and treatment-related outcomes in breast and prostate cancer patients

**Figure 1** and **Figure 2** show the IRs for the endocrine treatment prescriptions in breast and prostate cancer patients over the whole observation period. It is evident that prescriptions of AIs, tamoxifen, first generation ADT, GnRH agonists, and GnRH agonists with first generation ADT sharply reduced at the time of the first lockdown. **Figure 3** and **Figure 4** show the IRR of endocrine treatment prescriptions in breast and prostate cancer patients during the lockdown and post-lockdown periods compared to pre-pandemic rates. In patients with breast cancer, during the initial lockdown, prescriptions of AIs increased compared to the pre-pandemic period (IRR: 1.22 [95% Confidence Interval: 1.11-1.34]) and remained elevated across the majority of the post-lockdown periods. In patients with prostate cancer, during the initial lockdown, there was an increase in prescriptions of first-generation ADT compared to pre-lockdown (IRR: 1.23 [95% CI: 1.08-1.4) which remained elevated across the majority of the post-lockdown periods, and at the same time a decrease in prescriptions of GnRH agonists (IRR: 0.85 [95% CI: 0.76-0.95]). Rates remained below pre-pandemic rates for GnRH agonists until the third lockdown. First-generation ADT and GnRH agonists or antagonists, singularly or in combination, were more frequently prescribed from March 2021 onwards.

**Figure 1:**
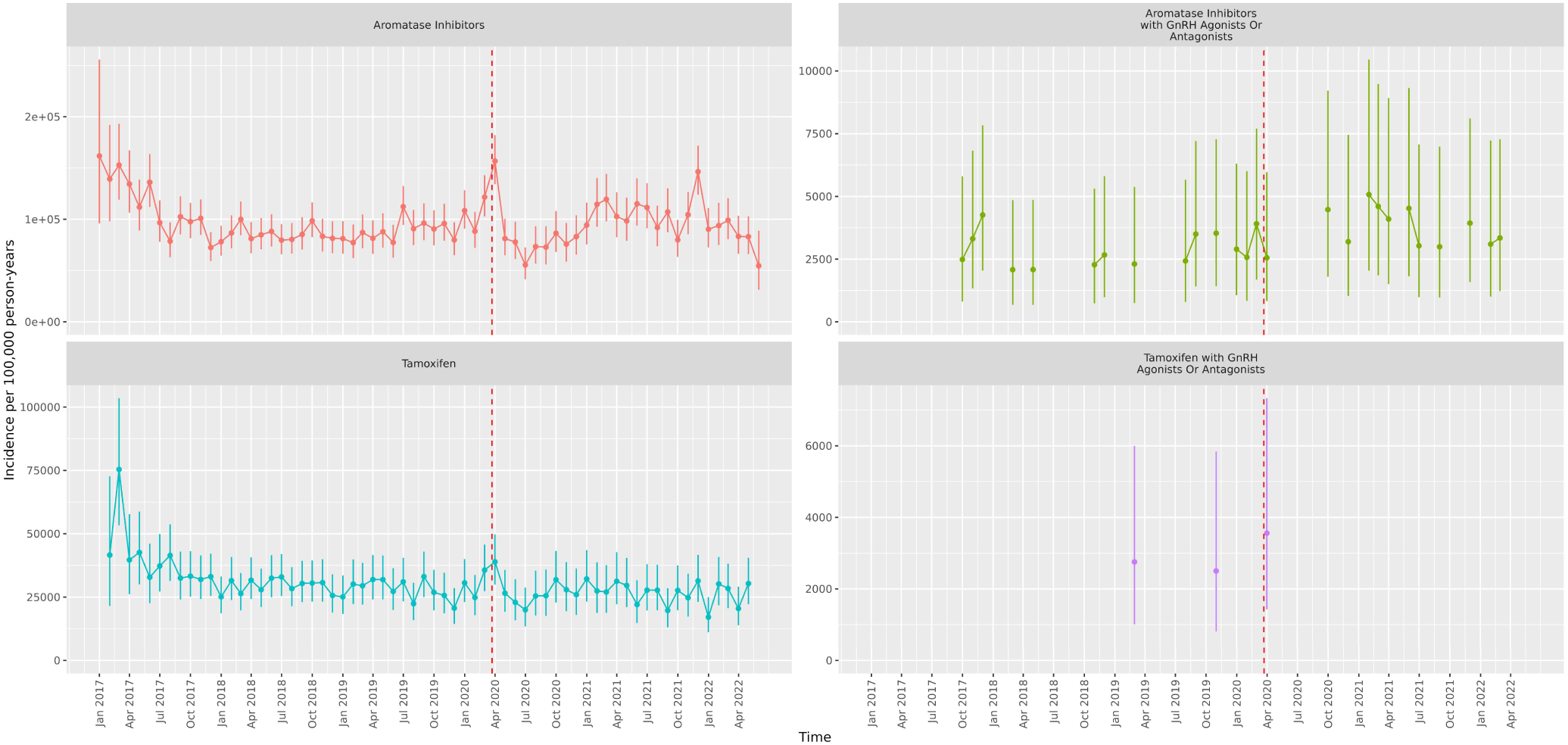
Incidence Rates (and 95% confidence intervals) of endocrine treatments in incident breast cancer patients from January 2017 to June 2022 **Note.** Dashed line indicates start of pandemic. Gaps between values indicate absence of data for the corresponding months.

**Figure 2:**
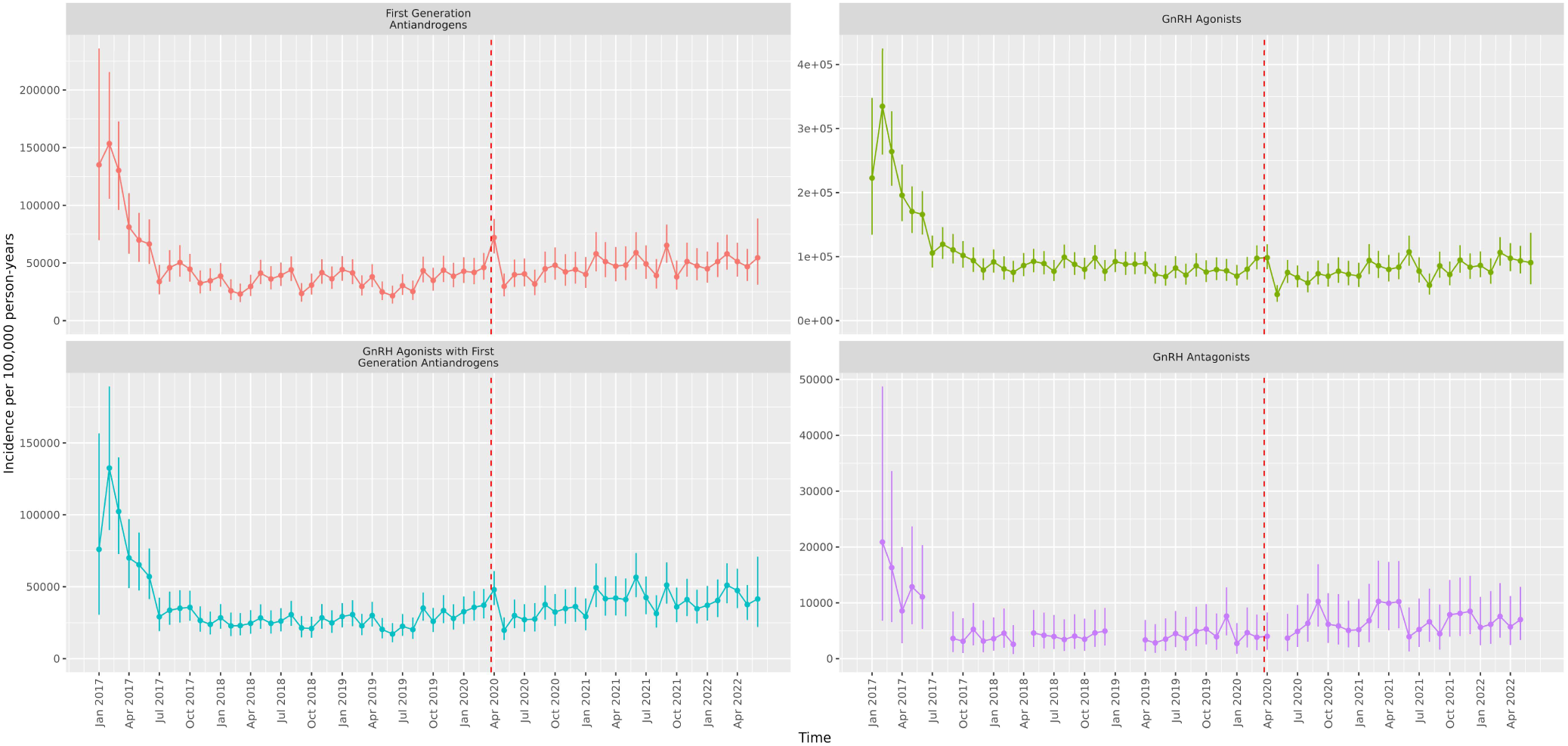
Incidence Rates (and 95% confidence intervals) of endocrine treatments in incident prostate cancer patients from January 2017 to June 2022 **Note.** Dashed line indicates start of pandemic. Gaps between values indicate absence of data for the corresponding months.

**Figure 3.**
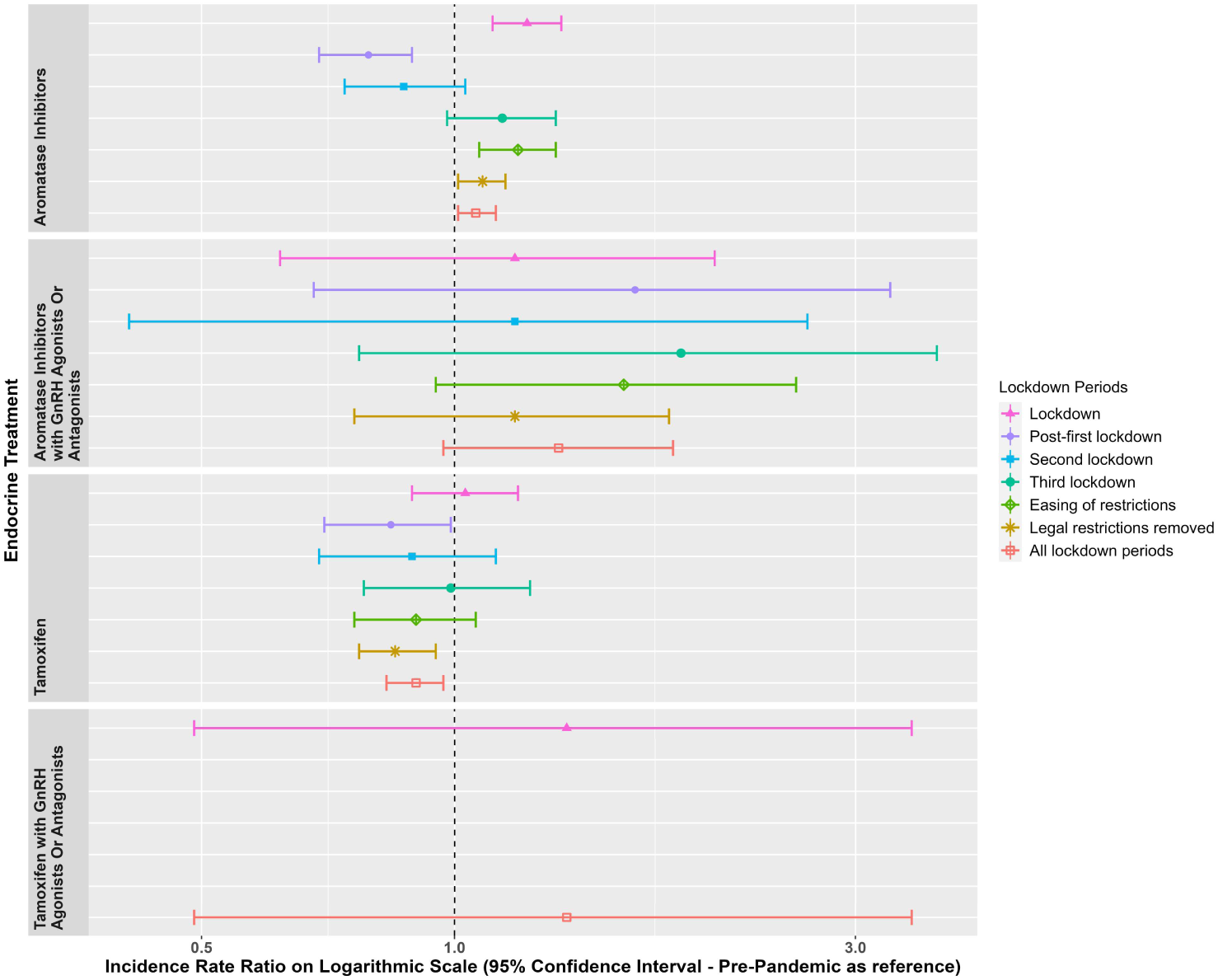
Incidence Rate Ratios (and 95% confidence intervals) of endocrine treatments in breast cancer patients in the post-lockdown periods compared to pre-pandemic rates. **Note.** Dashed line indicates start of pandemic. Lockdown periods defined as: Lockdown (March 2020 to June 2020); post-first lockdown (July 2020 to October 2020); second lockdown (Nov 2020 to Dec 2020); third lockdown (Jan 2021 to March 2021); easing of restrictions (April 2021 to June 2021); legal restrictions removed (July 2021 to June 2022); all lockdown periods (March 2020 to June 2022).

**Figure 4.**
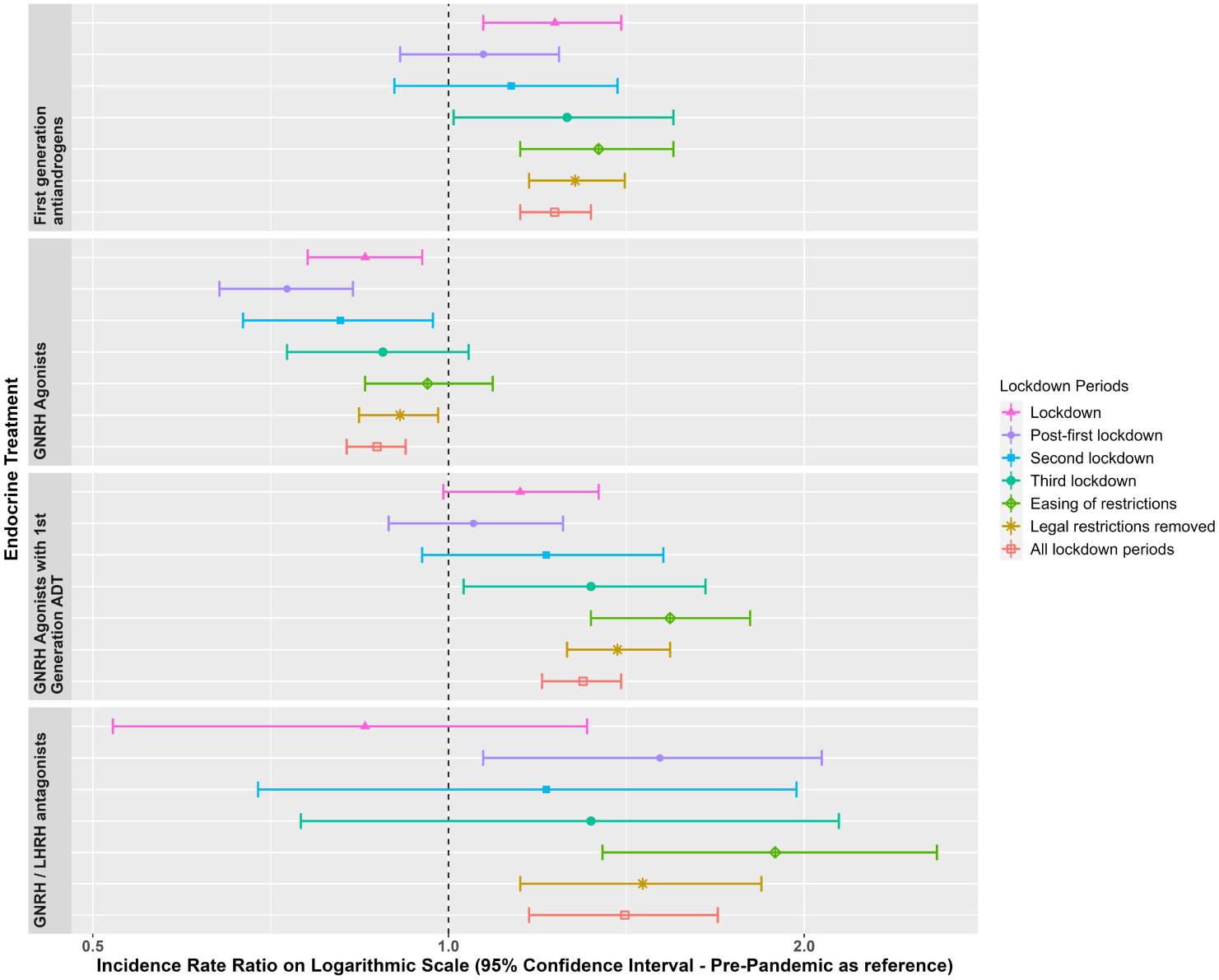
Incidence Rate Ratios (and 95% confidence intervals) of endocrine treatments in prostate cancer patients in the post-lockdown periods compared to pre-pandemic rates. **Note:** Dashed line indicates start of pandemic. Lockdown periods defined as: Lockdown (March 2020 to June 2020); post-first lockdown (July 2020 to October 2020); second lockdown (Nov 2020 to Dec 2020); third lockdown (Jan 2021 to March 2021); easing of restrictions (April 2021 to June 2021); legal restrictions removed (July 2021 to June 2022); all lockdown periods (March 2020 to June 2022).

IRR, number of events and IR which show the data used to derive Figures 1-4 are included in **Supplementary Tables S3 to S5** for breast cancer and **Supplementary Tables S6 to S8** for prostate cancer.

**Figure 5** and **Figure 6** show the IR of endocrine treatment-related outcomes in breast cancer patients on AIs and prostate cancer patients on any endocrine treatment over the whole observation period. It is evident that in breast cancer patients on AIs diagnoses of osteopenia and osteoporosis were not being made immediately following the first lockdown. There were no clear patterns for prostate cancer patients, largely due to small numbers of events. **Figure 7** and **Figure 8** show the IRR of endocrine treatment-related outcomes in breast cancer patients on AIs and prostate cancer patients on any endocrine treatment during the lockdown and post-lockdown periods compared to pre-pandemic rates. Prescriptions of bisphosphonates were significantly reduced across all lockdown periods between March 2020 and June 2022 (IRR range: 0.40-0.62) for breast cancer patients on AIs, as were diagnoses of osteopenia (IRR range: 0.31-0.6) and osteoporosis (all except for the post-first lockdown period) (IRR range: 0.36-0.55). For breast cancer patients on tamoxifen monthly counts of all treatment-related outcomes were too small to be included in IR analyses (counts per month <5). For prostate cancer patients on any endocrine treatments, IR were no different pre-pandemic compared to after March 2020 for bisphosphonates, and monthly counts too small for osteopenia and osteoporosis.

**Figure 5.**
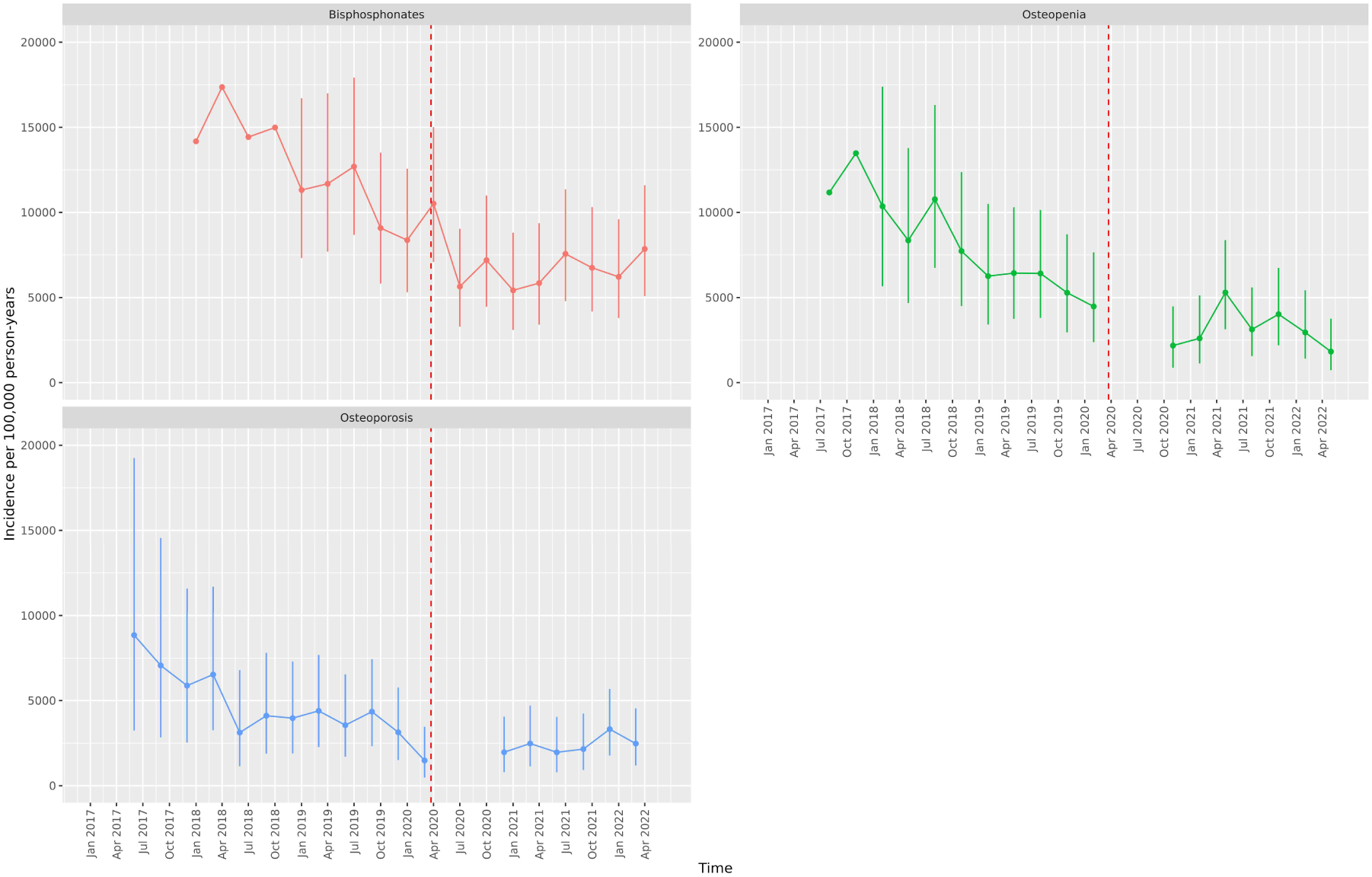
Incidence Rates (and 95% confidence intervals) of endocrine treatment-related outcomes in breast cancer patients on aromatase inhibitors from January 2017 to June 2022. **Note.** Dashed line indicates start of pandemic. Gaps between values indicate absence of data for the corresponding months.

**Figure 6.**
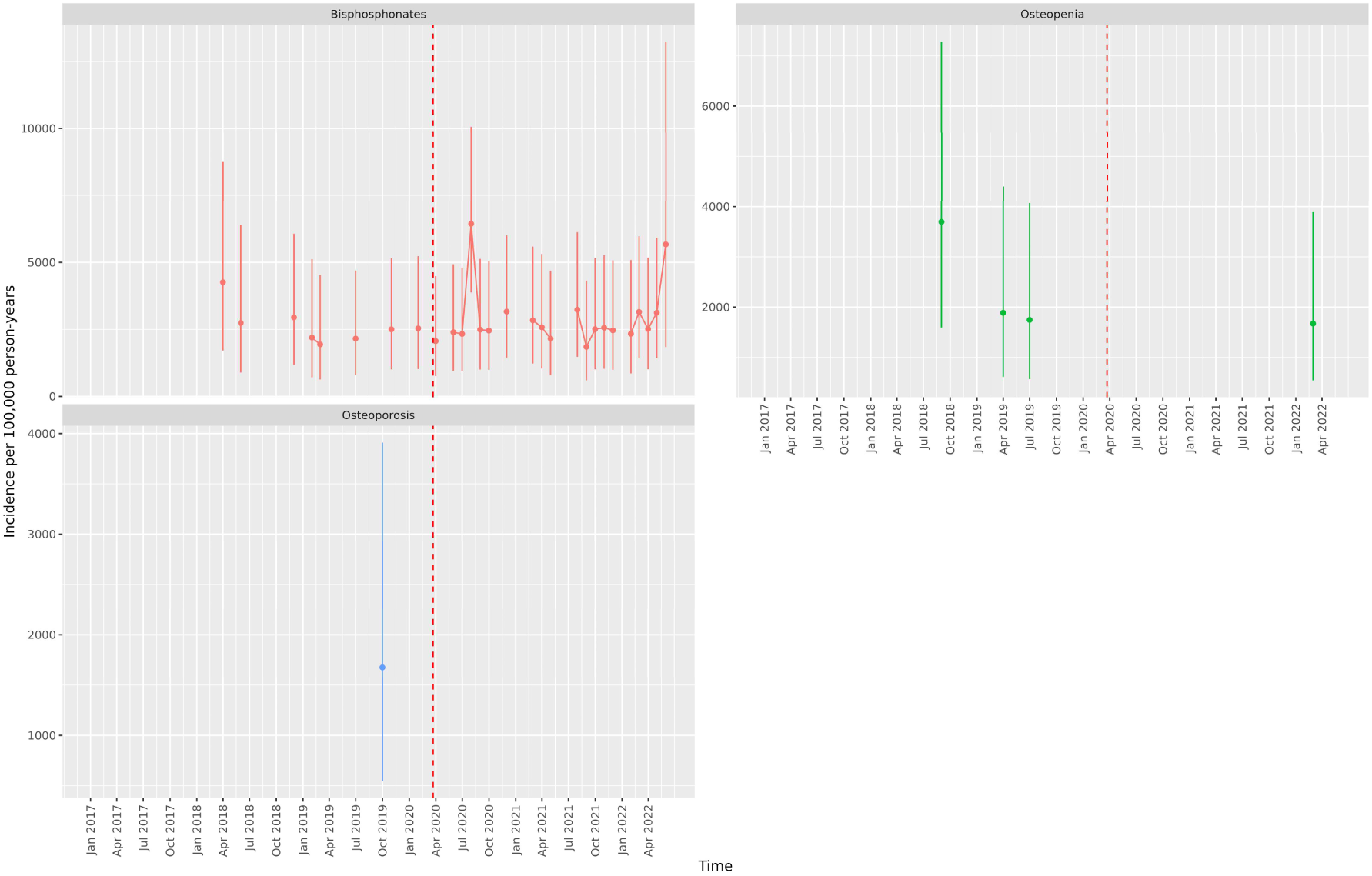
Incidence Rates (and 95% confidence intervals) of endocrine treatment-related outcomes in prostate cancer patients on endocrine treatments from January 2017 to June 2022. **Note:** Dashed line indicates start of pandemic. Gaps between values indicate absence of data for the corresponding months.

**Figure 7.**
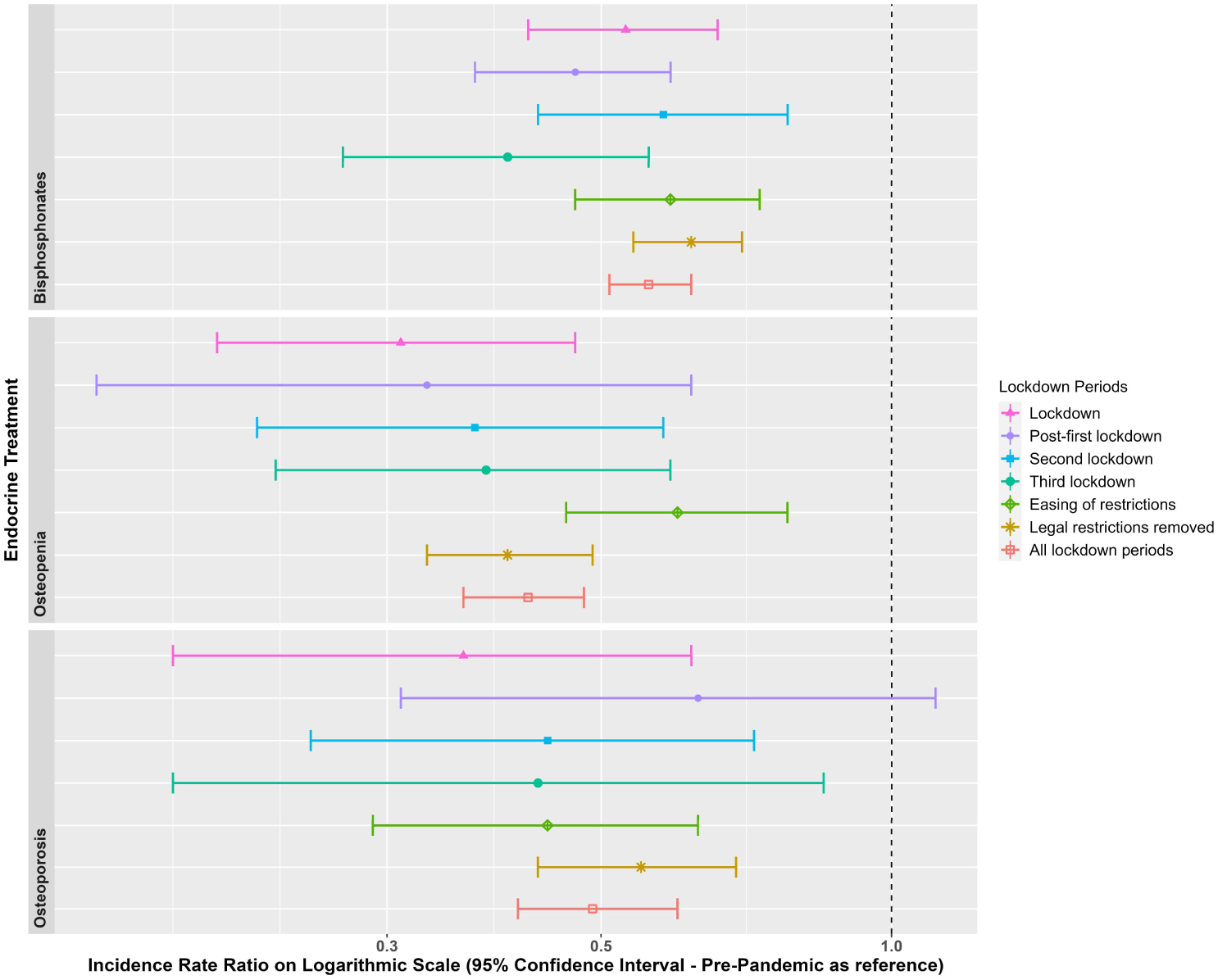
Incidence Rate Ratios (and 95% confidence intervals) of endocrine treatment-related outcomes in breast cancer patients on aromatase inhibitors in the extended post-lockdown periods compared to pre-pandemic rates. **Note:** Dashed line indicates start of pandemic. Lockdown periods defined as: Lockdown (March 2020 to June 2020); post-first lockdown (July 2020 to October 2020); second lockdown (Nov 2020 to Dec 2020); third lockdown (Jan 2021 to March 2021); easing of restrictions (April 2021 to June 2021); legal restrictions removed (July 2021 to June 2022); all lockdown periods (March 2020 to June 2022).

**Figure 8.**
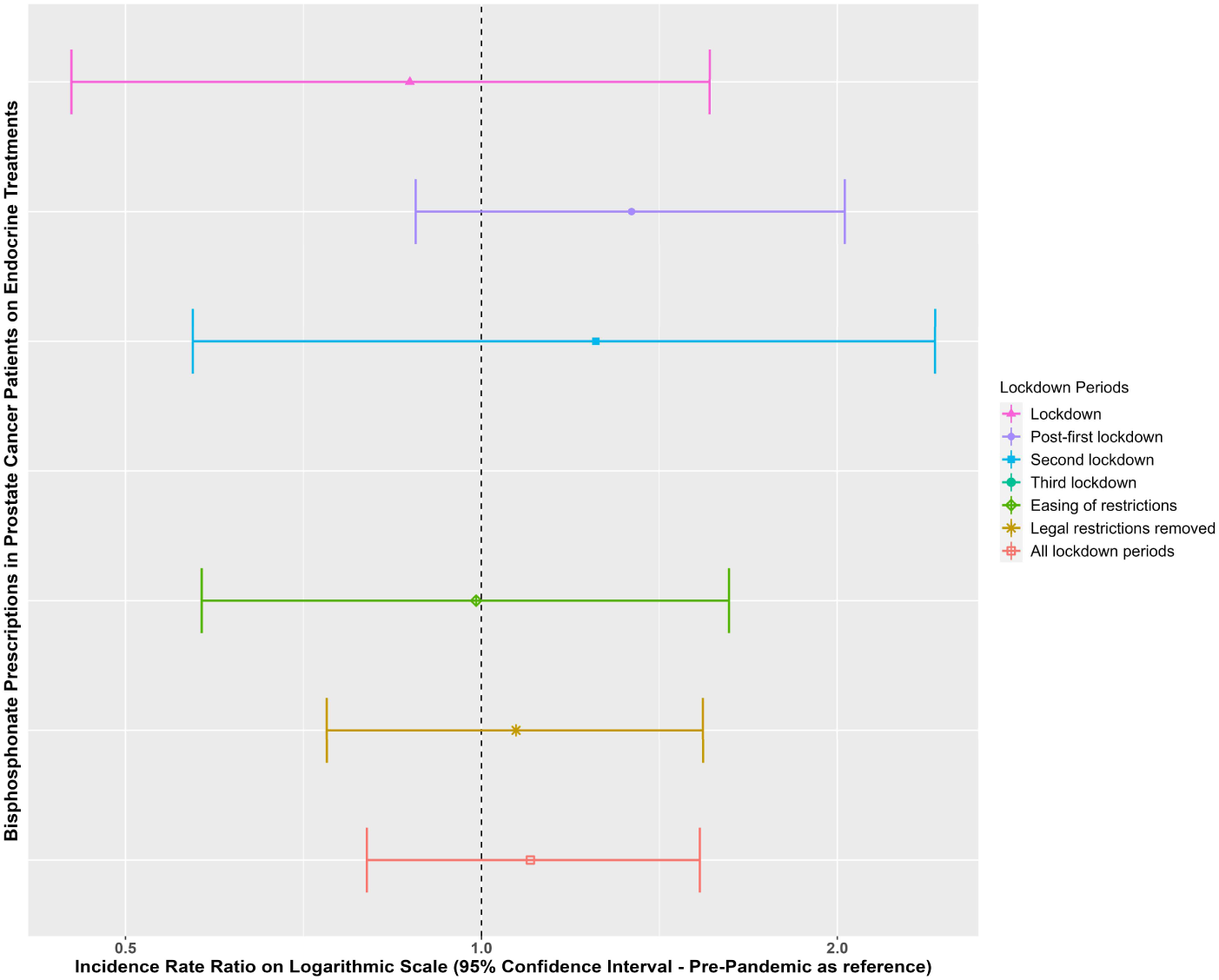
Incidence Rate Ratios (and 95% confidence intervals) of endocrine treatment-related outcomes (bisphosphonates) in prostate cancer patients on endocrine treatments in the extended post-lockdown periods compared to pre-pandemic rates. **Note:** Dashed line indicates start of pandemic. Lockdown periods defined as: Lockdown (March 2020 to June 2020); post-first lockdown (July 2020 to October 2020); second lockdown (Nov 2020 to Dec 2020); third lockdown (Jan 2021 to March 2021); easing of restrictions (April 2021 to June 2021); legal restrictions removed (July 2021 to June 2022); all lockdown periods (March 2020 to June 2022). Note that counts of prescriptions for third lockdown were <5 and so too small to calculate an incidence rate ratio.

IRR, number of events and IR which show the data used to derive Figures 5-8 are included in **Supplementary Tables S9 to S11** for breast cancer patients on AIs and **Supplementary Tables S12 to S14** for prostate cancer patients on any endocrine treatment.

## Discussion

In this study we examined the impact of the changing social restrictions imposed by the COVID-19 pandemic on the incidence and trends of endocrine treatments, and secondarily endocrine treatment-related outcomes of osteopenia and osteoporosis, and prescriptions of bisphosphonates, in breast and prostate cancer patients on endocrine treatments in the UK from January 2017 to June 2022.

In the months immediately following the first lockdown, incidence of prescriptions of AIs in breast cancer patients, and first-generation ADT in prostate cancer patients, increased compared to pre-pandemic rates, and remained elevated across the majority of the post-lockdown period between March 2020 and June 2022. This mirrors recommendations by some European guidelines for the management of breast and prostate cancer patients diagnosed during the early pandemic: delaying surgery or radiotherapy in the first 3-6 months of the pandemic and instead prescribing endocrine therapy [5]. Whilst delaying surgery or radiotherapy for breast or prostate cancer was not an official change to UK management guidelines, the results presented here demonstrate that approaches that limited in-patient hospital time appear to have been implemented in the UK during the pandemic (though it should be acknowledged that our results from primary care do not allow us to examine reductions in surgery or radiotherapy). This is in line with other research from the UK and worldwide. Indeed, one UK study demonstrated that alterations to breast cancer management were implemented in nearly 60% of patients, and many surgical interventions were substituted with ‘bridging’ endocrine therapy [12]. In the Netherlands neo-adjuvant endocrine therapies for breast cancer increased by 339% during lockdown [13]. As well as reduced availability of surgical resources, radiotherapies and hospital beds, concern that chemotherapy-induced immunosuppression would increase risk for COVID-19 complications may have influenced clinicians’ decisions to switch patients to alternative therapies [14]. An international survey of breast cancer management strategies indicated that 51% of clinician respondents reported modifications to chemotherapy treatments during the pandemic, and that 68% considered postponing surgery and administering endocrine treatments to patients with luminal A disease during the pandemic [14].

With regards changes to prostate cancer management, it is perhaps no surprise that prescriptions of GnRH analogues were reduced across the pandemic as these drugs are typically injected by a clinician, whereas the first-generation ADT therapies (including nilutamide, flutamide and bicalutamide) can be administered orally. That said, initial concerns about ADT increasing SARS-Cov-2 infection risk, COVID-19 complications requiring hospitalization, and mortality [15] might have led clinicians to be cautious about prescribing such medications in the early pandemic. Despite contradictory evidence, several systematic reviews and meta-analyses have now demonstrated no association between ADT and COVID-19 complications [16-19].

Whilst endocrine therapies can be effective in neoadjuvant settings for breast and prostate cancer, the use of some endocrine therapies has been associated with poor bone health. One study demonstrated that AIs exhibit a significant increased relative risk of 1.3 for bone loss (including osteopenia and osteoporosis) compared to patients not treated with AIs [20]. Likewise, the use of endocrine therapy in the treatment of prostate cancer has been shown to be associated with around 4.6% bone loss per year in men treated with GnRH analogues compared to a typical rate of 0.5% per year in healthy men [21]. In a small study of 105 patients treated with ADT for prostate cancer, prevalence of osteoporosis increased from around 10% at the beginning of the study to 22% at 2-year follow-up [22].

Given the increased use of AIs in breast cancer patients and ADT in prostate cancer patients across the pandemic, a secondary aim of this study was to investigate the rate of diagnosis of secondary diseases such as osteoporosis before and after lockdown, and the possibility that such diagnoses may have been missed due to poorer treatment evaluation during the pandemic for these two therapies. Our results indicate that diagnoses of osteopenia and osteoporosis were reduced across the pandemic compared to the pre-pandemic era for new AI users. This is likely driven by delayed assessments, bone scans and palliative treatment with bisphosphonates during the pandemic. Indeed, in a worldwide survey to primarily medical oncologists, 64% of respondents reported reduced frequency of DEXA scans in the first four months of the pandemic, and difficulties with access to General Practitioner (GP) or hospital-administered treatments such as intravenous bisphosphonates or subcutaneous denosumab [23]. Sixty-six percent of respondents reported that adjuvant intravenous bisphosphonate use had been impacted by the pandemic, in terms of delayed treatment, missed appointments, and reduced clinical capacity, requiring a switch from intravenous to oral administration; whilst nearly a quarter of respondents reported decreased use of oral bisphosphonates. This is in line with our results which show that prescriptions of bisphosphonates were indeed reduced across the pandemic for breast cancer patients on AIs (though it should be noted that this pattern is seen in other populations, not limited to cancer patients [24]).

In contrast, no differences in bone-related treatment outcomes across the lockdown periods were observed for new prostate cancer ADT-users. This could be explained by the fact that bone health assessments are less common in the male population compared to females (whose risk for bone related complications, particularly after menopause is higher than males [25]). Alternative explanations include the fact that first-generation ADT used as monotherapy (such as bicalutamide) preserve bone mineral density, reducing the likelihood of bone-related complications. In contrast, GnRH agonists do affect bone health, and the decreased prescriptions observed during the pandemic may have consequently reduced any pandemic-related effects on bone-related outcomes in this population.

### Strengths and Weaknesses of the Study

This study benefits from the strengths of CPRD GOLD, known for its extensive UK population coverage and comprehensive healthcare records [8], facilitating thorough phenotyping of breast and prostate cancer, as well as endocrine treatments. The longitudinal nature of the database enabled an examination of the trend in endocrine prescriptions over a period of nearly 2 years beyond the start of the pandemic. However, this study also has some limitations. First, as these data are derived from primary care and not linked to cancer registry or inpatient data, we were unable to investigate the hypothesis that endocrine treatments may have increased in use across the pandemic because of delays in surgery, radiotherapy and other hospital-initiated treatments. Furthermore, our assessment of rates of endocrine therapies that may be administered in secondary care (e.g. GnRH analogues) may be underestimated given our focus on primary care data. Further research on secondary care, hospital settings and cancer registries is therefore needed. Second, the generalizability of findings is predominantly limited to England and Scotland in these patient cohorts, with less representation from Wales and Northern Ireland. That said, the composition of patients and practices in the database have changed over time. Indeed, with the advent of the CPRD AURUM database, some practices were transferred out of GOLD and into AURUM, and so across the time period of this study, practices from England and Northern Ireland reduced, whilst practices from Scotland and Wales increased. Reassuringly, the IR of the endocrine treatments in the breast and prostate cancer populations in the three broad time-periods across regions were largely similar, except for smaller counts of AIs with GnRH and Tamoxifen with GnRH in England and Northern Ireland post-lockdown, and slightly higher rates of GnRH agonists with first generation ADT post-lockdown, likely reflecting the change in population composition (see **supplementary Figures S1 and S2**).

### Conclusions

During the early months of the pandemic, newly diagnosed breast cancer or prostate cancer patients were more likely to be prescribed AIs (for breast), or first-generation antiandrogens (for prostate cancer), compared to before the pandemic. This is likely driven by delays in surgery, radiotherapy or other treatments requiring hospital visits, and endocrine therapy being prescribed as a neoadjuvant/bridging therapy. Despite this initial increased prescribing of aromatase inhibitors in breast cancer patients, these patients received fewer prescriptions of bisphosphonates to protect against bone thinning as a result of endocrine exposure. At the same time, diagnosis rates of osteopenia and osteoporosis were reduced compared to pre-pandemic, potentially due to the lack of diagnostic testing for these conditions during the pandemic. These results highlight the need to follow-up breast cancer patients on aromatase inhibitors in the coming years for signs of bone thinning, given the relatively poorer management of endocrine treatment-related side-effects in this population during the pandemic.

## Data Availability

This study is based on data from the Clinical Practice Research Datalink (CPRD) obtained under licence from the UK Medicines and Healthcare products Regulatory Agency. The data is provided by patients and collected by the NHS as part of their care and support. The interpretation and conclusions contained in this study are those of the author/s alone. Patient- level data used in this study was obtained through an approved application to the CPRD RDG process (application number 22_002331). Data is only available directly from CPRD following RDG approval Details on how to apply for data access can be found at https://cprd.com/data-access. Analytical code and code lists for identifying the events are available in GitHub repositories: https://github.com/oxford-pharmacoepi/CancerCovidEndocrineTx

## Additional Information

## Acknowledgements

None.

## Authors’ contributions

MPM, MCS, AJ, DN, NLB and DPA conceived the study. MPM, MCS, DPA and NB were involved in developing the study design. NLB, MCS and MPM wrote and executed the statistical code. AD and WYM mapped the CPRD GOLD data to the OMOP CDM. ARS provided clinical expertise on phenotype definitions. NLB wrote the manuscript. All authors interpreted the results. All authors were involved in the editing of drafts of the manuscript. All authors approved the final version and had final responsibility for the decision to submit for publication. DPA is the guarantor. The corresponding author attests that all listed authors meet authorship criteria and that no others meeting the criteria have been omitted.

## Ethics approval

The protocol for this research was approved by the Research Data Governance (RDG) Board of the Medicine and Healthcare products Regulatory Agency database research (protocol number 22_002331).

## Data availability

This study is based on data from the Clinical Practice Research Datalink (CPRD) obtained under licence from the UK Medicines and Healthcare products Regulatory Agency. The data is provided by patients and collected by the NHS as part of their care and support. The interpretation and conclusions contained in this study are those of the author/s alone. Patient-level data used in this study was obtained through an approved application to the CPRD RDG process (application number 22_002331). Data is only available directly from CPRD following RDG approval Details on how to apply for data access can be found at https://cprd.com/data-access. Analytical code and code lists for identifying the events are available in GitHub repositories: https://github.com/oxford-pharmacoepi/CancerCovidEndocrineTx

