## Supplementary material for "Collateral effects of the COVID-19 pandemic on endocrine treatments for breast and prostate cancer in the UK: implications for bone health"

### Supplementary Tables and Figures

**Table S1.** Breast cancer patient starting counts, final counts after applying excision criteria, and reasons for exclusion

| Denominator count (n) | Reason for exclusion | Excluded (n) |
|---|---|---|
| 109,498 | Starting population | NA |
| 109,498 | Missing year of birth | 0 |
| 109,498 | Missing sex | 0 |
| 109,498 | Cannot satisfy age criteria during the study period based on year of birth | 0 |
| 18,017 | No observation time available during study period (January 2017 to December 2021) | 91,481 |
| 18,017 | Doesn't satisfy age criteria during the study period | 0 |
| 17,765 | Prior history requirement not fulfilled during study period | 252 |
| 13,774 | No observation time available after applying age, prior history and, if applicable, strata criteria | 3,991 |
| 13,760 | Excluded due to prior breast cancer diagnosis (do not pass outcome washout during study period) | 14 |

**Table S2.** Prostate cancer patient starting counts, final counts after applying excision criteria, and reasons for exclusion

| Denominator count (n) | Reason for exclusion | Excluded (n) |
|---|---|---|
| 78,911 | Starting population | NA |
| 78,911 | Missing year of birth | 0 |
| 78,911 | Missing sex | 0 |
| 78,911 | Cannot satisfy age criteria during the study period based on year of birth | 0 |
| 15,433 | No observation time available during study period (January 2017 to December 2021) | 63,478 |
| 15,433 | Doesn't satisfy age criteria during the study period | 0 |
| 15,234 | Prior history requirement not fulfilled during study period | 199 |
| 15,230 | Not male | 4 |
| 8,821 | No observation time available after applying age, prior history and, if applicable, strata criteria | 6,425 |
| 8,805 | Excluded due to prior prostate cancer diagnosis (do not pass outcome washout during study period) | 0 |

**Table S3.** Incidence Rate Ratios (95% confidence intervals) of endocrine treatments in breast cancer patients

| **Endocrine Treatment** | **Lockdown (March 2020-June 2020)** | **Post-first lockdown (July 2020-Oct 2020)** | **Second lockdown (Nov 2020-Dec 2020)** | **Third lockdown (Jan 2021-Feb 2021)** | **Easing of restrictions (March 2021-June 2021** | **Legal restrictions removed (July 2021-June 2022)** | **All lockdown periods (March 2020-June 2022)** |
|---|---|---|---|---|---|---|---|
| Aromatase Inhibitors | 1.22 (1.11 to 1.34) | 0.79 (0.69 to 0.89) | 0.87 (0.74 to 1.03) | 1.14 (0.98 to 1.32) | 1.19 (1.08 to 1.32) | 1.08 (1.01 to 1.15) | 1.06 (1.01 to 1.12) |
| Aromatase Inhibitors with GnRH Agonists Or Antagonists | 1.18 (0.62 to 2.04) | 1.64 (0.68 to 3.31) | 1.18 (0.41 to 2.63) | 1.86 (0.77 to 3.75) | 1.59 (0.95 to 2.55) | 1.18 (0.76 to 1.8) | 1.33 (0.97 to 1.82) |
| Tamoxifen | 1.03 (0.89 to 1.19) | 0.84 (0.7 to 0.99) | 0.89 (0.69 to 1.12) | 0.99 (0.78 to 1.23) | 0.9 (0.76 to 1.07) | 0.85 (0.77 to 0.95) | 0.9 (0.83 to 0.97) |
| Tamoxifen with GnRH Agonists Or Antagonists | 1.36 (0.49 to 3.5) | NA | NA | NA | NA | NA | 1.36 (0.49 to 3.5) |

Note. NA = counts <5 are not reported.

| **Table S4.** Number of events and person months (in parentheses) of endocrine treatment-related outcomes in breast cancer patients | | | | | | | | |
|---|---|---|---|---|---|---|---|---|
| **Endocrine Treatment-Related Outcome** | **Pre-COVID (Jan 2017-Feb 2020)** | **Lockdown (March 2020-June 2020)** | **Post-first lockdown (July 2020-Oct 2020)** | **Second lockdown (Nov 2020-Dec 2020)** | **Third lockdown (Jan 2021-Feb 2021)** | **Easing of restrictions (March 2021-June 2021** | **Legal restrictions removed (July 2021-June 2022)** | **All lockdown periods (March 2020-June 2022)** |
| Aromatase Inhibitors | 4095 (53825.84) | 479 (5177.46) | 260 (4349.73) | 143 (2156.85) | 181 (2091.56) | 389 (4283.66) | 1087 (13291.33) | 2539 (31350.6) |
| Aromatase Inhibitors with GnRH Agonists Or Antagonists | 78 (33543.39) | 13 (4803.29) | 7 (1878.24) | 5 (1878.24) | 7 (1655.75) | 20 (5439.31) | 28 (10207.9) | 80 (25862.74) |
| Tamoxifen | 1833 (72253.17) | 203 (7785.56) | 138 (6485.16) | 69 (3076.21) | 74 (2965.42) | 142 (6214.14) | 421 (19472.07) | 1047 (45998.55) |
| Tamoxifen with GnRH Agonists Or Antagonists | 11 (5011.71) | 7 (2360.64) | NA | NA | NA | NA | NA | 7 (2360.64) |

Note. NA = counts <5 are not reported.

**Table S5.** Incidence Rates (95% confidence intervals) per 100,000 person months of endocrine treatments in breast cancer patients

| Endocrine Treatment | Pre-COVID (Jan 2017-Feb 2020) | Lockdown (March 2020-June 2020) | Post-first lockdown (July 2020-Oct 2020) | Second lockdown (Nov 2020-Dec 2020) | Third lockdown (Jan 2021-Feb 2021) | Easing of restrictions (March 2021-June 2021 | Legal restrictions removed (July 2021-June 2022) | All lockdown periods (March 2020-June 2022) |
|---|---|---|---|---|---|---|---|---|
| Aromatase Inhibitors | 7607.9 (7376.6 to 7844.5) | 9251.6 (8441.6 to 10118.5) | 5977.4 (5272.8 to 6749.8) | 6630.1 (5587.9 to 7810.1) | 8653.8 (7439 to 10010.5) | 9081 (8200.9 to 10029.8) | 8178.3 (7699.2 to 8679.3) | 8098.7 (7786.7 to 8420) |
| Aromatase Inhibitors with GnRH Agonists Or Antagonists | 232.5 (183.8 to 290.2) | 270.6 (144.1 to 462.8) | 372.7 (149.8 to 767.9) | 266.2 (86.4 to 621.2) | 422.8 (170 to 871.1) | 367.7 (224.6 to 567.9) | 274.3 (182.3 to 396.4) | 309.3 (245.3 to 385) |
| Tamoxifen | 2536.9 (2422.1 to 2655.8) | 2607.4 (2261 to 2991.8) | 2127.9 (1787.7 to 2514) | 2243 (1745.2 to 2838.7) | 2495.4 (1959.4 to 3132.8) | 2285.1 (1924.7 to 2693.4) | 2162.1 (1960.5 to 2378.8) | 2276.2 (2140.4 to 2418.3) |
| Tamoxifen with GnRH Agonists Or Antagonists | 219.5 (109.6 to 392.7) | 296.5 (119.2 to 611) | NA | NA | NA | NA | NA | 296.5 (119.2 to 611) |

Note. NA = counts <5 are not reported.

**Table S6.** Incidence Rate Ratios (95% confidence intervals) of endocrine treatments in prostate cancer patients

| Endocrine Treatment | Lockdown (March 2020-June 2020) | Post-first lockdown (July 2020-Oct 2020) | Second lockdown (Nov 2020-Dec 2020) | Third lockdown (Jan 2021-Feb 2021) | Easing of restrictions (March 2021-June 2021 | Legal restrictions removed (July 2021-June 2022) | All lockdown periods (March 2020-June 2022) |
|---|---|---|---|---|---|---|---|
| First generation antiandrogens | 1.23 (1.07 to 1.4) | 1.07 (0.91 to 1.24) | 1.13 (0.9 to 1.39) | 1.26 (1.01 to 1.55) | 1.34 (1.15 to 1.55) | 1.28 (1.17 to 1.41) | 1.23 (1.15 to 1.32) |
| GNRH Agonists | 0.85 (0.76 to 0.95) | 0.73 (0.64 to 0.83) | 0.81 (0.67 to 0.97) | 0.88 (0.73 to 1.04) | 0.96 (0.85 to 1.09) | 0.91 (0.84 to 0.98) | 0.87 (0.82 to 0.92) |
| GNRH Agonists with 1st Generation ADT | 1.15 (0.99 to 1.34) | 1.05 (0.89 to 1.25) | 1.21 (0.95 to 1.52) | 1.32 (1.03 to 1.65) | 1.54 (1.32 to 1.8) | 1.39 (1.26 to 1.54) | 1.30 (1.20 to 1.40) |
| GNRH / LHRH antagonists | 0.85 (0.52 to 1.31) | 1.51 (1.07 to 2.07) | 1.21 (0.69 to 1.97) | 1.32 (0.75 to 2.14) | 1.89 (1.35 to 2.59) | 1.46 (1.15 to 1.84) | 1.41 (1.17 to 1.69) |

**Table S7.** Number of events and person months (in parentheses) of treatment related outcomes in prostate cancer patients

| Endocrine Treatment | Pre-COVID (Jan 2017-Feb 2020) | Lockdown (March 2020-June 2020) | Post-first lockdown (July 2020-Oct 2020) | Second lockdown (Nov 2020-Dec 2020) | Third lockdown (Jan 2021-Feb 2021) | Easing of restrictions (March 2021-June 2021 | Legal restrictions removed (July 2021-June 2022) | All lockdown periods (March 2020-June 2022) |
|---|---|---|---|---|---|---|---|---|
| First generation antiandrogens | 1880 (58541.7) | 245 (6221.31) | 177 (5164.98) | 83 (2298.32) | 86 (2128.39) | 189 (4407.66) | 555 (13471.97) | 1335 (33692.62) |
| GNRH Agonists | 3387 (43846.87) | 326 (4975.21) | 241 (4301.14) | 121 (1940.44) | 122 (1807.15) | 280 (3763.09) | 804 (11411.61) | 1894 (28198.64) |
| GNRH Agonists with 1st Generation ADT | 1525 (62216.54) | 187 (6614.97) | 145 (5614.26) | 74 (2499.02) | 74 (2298.78) | 178 (4703.61) | 486 (14233.69) | 1144 (35964.32) |
| GNRH / LHRH antagonists | 237 (62492.58) | 20 (6240.1) | 42 (7366.08) | 15 (3300.8) | 15 (3028.93) | 43 (6021.91) | 101 (18268.02) | 236 (44225.84) |

**Table S8.** Incidence Rates (95% confidence intervals) per 100,000 person months of endocrine treatments in prostate cancer patients

| Endocrine Treatment | Pre-COVID (Jan 2017-Feb 2020) | Lockdown (March 2020-June 2020) | Post-first lockdown (July 2020-Oct 2020) | Second lockdown (Nov 2020-Dec 2020) | Third lockdown (Jan 2021-Feb 2021) | Easing of restrictions (March 2021-June 2021 | Legal restrictions removed (July 2021-June 2022) | All lockdown periods (March 2020-June 2022) |
|---|---|---|---|---|---|---|---|---|
| First generation antiandrogens | 3211.4 (3067.8 to 3359.9) | 3938.1 (3460.4 to 4463.3) | 3426.9 (2940.7 to 3970.6) | 3611.3 (2876.4 to 4476.8) | 4040.6 (3232 to 4990.1) | 4288 (3698.4 to 4944.8) | 4119.7 (3784 to 4477.1) | 3962.3 (3752.6 to 4180.7) |
| GNRH Agonists | 7724.6 (7466.6 to 7989.2) | 6552.5 (5860.4 to 7303.8) | 5603.2 (4918 to 6357) | 6235.7 (5174.2 to 7450.9) | 6751 (5606.3 to 8060.7) | 7440.7 (6594.6 to 8365.2) | 7045.5 (6566.8 to 7549.8) | 6716.6 (6417.5 to 7026.1) |
| GNRH Agonists with 1st Generation ADT | 2451.1 (2329.6 to 2577.3) | 2826.9 (2436.3 to 3262.4) | 2582.7 (2179.4 to 3039) | 2961.2 (2325.1 to 3717.5) | 3219.1 (2527.7 to 4041.3) | 3784.3 (3248.8 to 4382.9) | 3414.4 (3117.6 to 3731.9) | 3180.9 (2999.3 to 3370.7) |
| GNRH / LHRH antagonists | 379.2 (332.5 to 430.7) | 320.5 (195.8 to 495) | 570.2 (410.9 to 770.7) | 454.4 (254.3 to 749.5) | 495.2 (277.2 to 816.8) | 714.1 (516.8 to 961.8) | 552.9 (450.3 to 671.8) | 533.6 (467.7 to 606.2) |

**Table S9.** Incidence Rate Ratios (95% confidence intervals) of endocrine treatment-related outcomes in breast cancer patients on aromatase inhibitors

| Endocrine Treatment | Lockdown | Post-first lockdown | Second lockdown | Third lockdown | Easing of restrictions | Legal restrictions removed | All lockdown periods (March 2020-June 2022) |
|---|---|---|---|---|---|---|---|
| Bisphosphonates | 0.53 (0.42 to 0.66) | 0.47 (0.37 to 0.59) | 0.58 (0.43 to 0.78) | 0.4 (0.27 to 0.56) | 0.59 (0.47 to 0.73) | 0.62 (0.54 to 0.7) | 0.56 (0.51 to 0.62) |
| Osteopenia | 0.31 (0.2 to 0.47) | 0.33 (0.15 to 0.62) | 0.37 (0.22 to 0.58) | 0.38 (0.23 to 0.59) | 0.6 (0.46 to 0.78) | 0.4 (0.33 to 0.49) | 0.42 (0.36 to 0.48) |
| Osteoporosis | 0.36 (0.18 to 0.62) | 0.63 (0.31 to 1.11) | 0.44 (0.25 to 0.72) | 0.43 (0.18 to 0.85) | 0.44 (0.29 to 0.63) | 0.55 (0.43 to 0.69) | 0.49 (0.41 to 0.60) |

| **Table S10.** Number of events and person months (in parentheses) of treatment related outcomes in breast cancer patients on aromatase inhibitors | | | | | | | |
|---|---|---|---|---|---|---|---|
| **Endocrine Treatment-Related Outcome** | **Pre-COVID** | **Lockdown** | **Post-first lockdown** | **Second lockdown** | **Third lockdown** | **Easing of restrictions** | **Legal restrictions removed** | **All lockdown periods (March 2020-June 2022)** |
| Bisphosphonates | 845 (75158.47) | 83 (13982.06) | 74 (14113.48) | 45 (6890.61) | 30 (6761.46) | 94 (14155.56) | 294 (42448) | 620 (98351.18) |
| Osteopenia | 502 (81468.39) | 22 (11536.2) | 8 (3993.59) | 18 (7893.26) | 18 (7758.52) | 60 (16160.16) | 117 (47476.27) | 243 (94818) |
| Osteoporosis | 312 (81262.78) | 11 (8089.92) | 10 (4221.21) | 14 (8354.43) | 7 (4302.85) | 29 (17259.73) | 87 (41219.78) | 158 (83447.92) |

**Table S11.** Incidence Rates (95% confidence intervals) per 100,000 person months of endocrine treatment-related outcomes in breast cancer patients on aromatase inhibitors

| Endocrine Treatment-related outcome | Pre-COVID (Jan 2017-Feb 2020) | Lockdown (March 2020-June 2020) | Post-first lockdown (July 2020-Oct 2020) | Second lockdown (Nov 2020-Dec 2020) | Third lockdown (Jan 2021-Feb 2021) | Easing of restrictions (March 2021-June 2021 | Legal restrictions removed (July 2021-June 2022) | All lockdown periods (March 2020-June 2022) |
|---|---|---|---|---|---|---|---|---|
| Bisphosphonates | 1124.3 (1049.8 to 1202.7) | 593.6 (472.8 to 735.9) | 524.3 (411.7 to 658.2) | 653.1 (476.3 to 873.8) | 443.7 (299.4 to 633.4) | 664 (536.6 to 812.6) | 692.6 (615.7 to 776.5) | 630.4 (581.7 to 682) |
| Osteopenia | 616.2 (563.5 to 672.5) | 190.7 (119.5 to 288.7) | 200.3 (86.5 to 394.7) | 228 (135.2 to 360.4) | 232 (137.5 to 366.7) | 371.3 (283.3 to 477.9) | 246.4 (203.8 to 295.4) | 256.3 (225.1 to 290.6) |
| Osteoporosis | 383.9 (342.5 to 429) | 136 (67.9 to 243.3) | 236.9 (113.6 to 435.7) | 167.6 (91.6 to 281.2) | 162.7 (65.4 to 335.2) | 168 (112.5 to 241.3) | 211.1 (169.1 to 260.3) | 189.3 (161 to 221.3) |

**Table S12.** Incidence Rate Ratios (95% confidence intervals) of endocrine treatment-related outcomes in prostate cancer population on any endocrine treatm

| Endocrine Treatment-related outcome | Lockdown | Post-first lockdown | Second lockdown | Third lockdown | Easing of restrictions | Legal restrictions removed | All lockdown periods (March 2020-June 2022) |
|---|---|---|---|---|---|---|---|
| Bisphosphonates | 0.87 (0.45 to 1.56) | 1.34 (0.88 to 2.03) | 1.25 (0.57 to 2.42) | NA | 0.99 (0.58 to 1.62) | 1.07 (0.74 to 1.54) | 1.1 (0.80 to 1.53) |

Note. NA = counts <5 are not reported.

**Table S13.** Number of events and person months (in parentheses) of endocrine treatment-related outcomes in prostate cancer patients on endocrine treatments

| Endocrine Treatment-Related Outcome | Pre-COVID | Lockdown | Post-first lockdown | Second lockdown | Third lockdown | Easing of restrictions | Legal restrictions removed | All lockdown periods (March 2020-June 2022) |
|---|---|---|---|---|---|---|---|---|
| Bisphosphonates | 49 (22849.87) | 13 (7002.61) | 40 (13941.52) | 9 (3413.45) | NA | 21 (9984.49) | 71 (31006.949) | 154 (65349.03) |

Note. NA = counts <5 are not reported.

**Table S14.** Incidence Rates (95% confidence intervals) per 100,000 person months of endocrine treatment-related outcomes in prostate cancer patients on any endocrine treatment

| Endocrine-Treatment related Outcome | Pre-COVID (Jan 2017-Feb 2020) | Lockdown (March 2020-June 2020) | Post-first lockdown 1 (July 2020-Oct 2020) | Second lockdown (Nov 2020-Dec 2020) | Third lockdown (Jan 2021-Feb 2021) | Easing of restrictions (March 2021-June 2021 | Legal restrictions removed (July 2021-June 2022) | All lockdown periods (March 2020-June 2022) |
|---|---|---|---|---|---|---|---|---|
| Bisphosphonates | 214.4 (158.6 to 283.5) | 185.6 (98.8 to 317.5) | 286.9 (205 to 390.7) | 263.7 (120.6 to 500.5) | NA | 210.3 (130.2 to 321.5) | 229 (178.8 to 288.8) | 235.7 (199.9 to 276) |
| Osteopenia | 195.3 (115.7 to 308.6) | NA | NA | NA | NA | NA | 139.4(45.3 to 325.3) | 139.4 (45.3 to 325.3) |
| Osteoporosis | 139.6 (45.3 to 325.8) | NA | NA | NA | NA | NA | NA | NA |

Note. NA = counts <5 are not reported.

**Figure S1.** Incidence Rates per 100,000 Person-Years (with 95% confidence intervals) of Endocrine Treatments in Breast Cancer Patients Before, During and After COVID-19 Lockdown Stratified by UK Region.

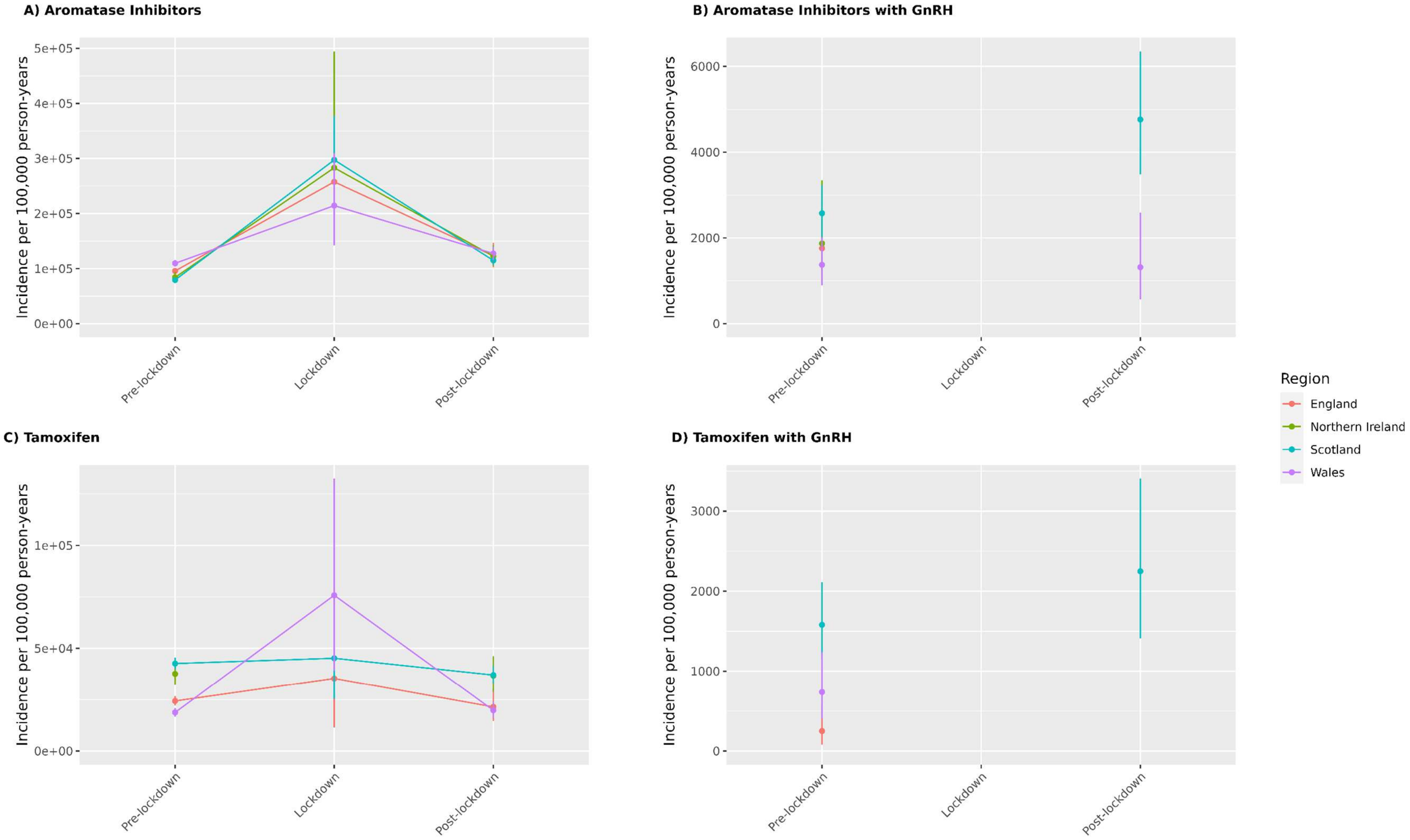


**Note.** GnRH = Gonadotrophin releasing hormone.

**Figure S2.** Incidence Rates per 100,000 Person-Years (with 95% confidence intervals) of Endocrine Treatments in Prostate Cancer Patients Before, During and After COVID-19 Lockdown Stratified by UK Region.

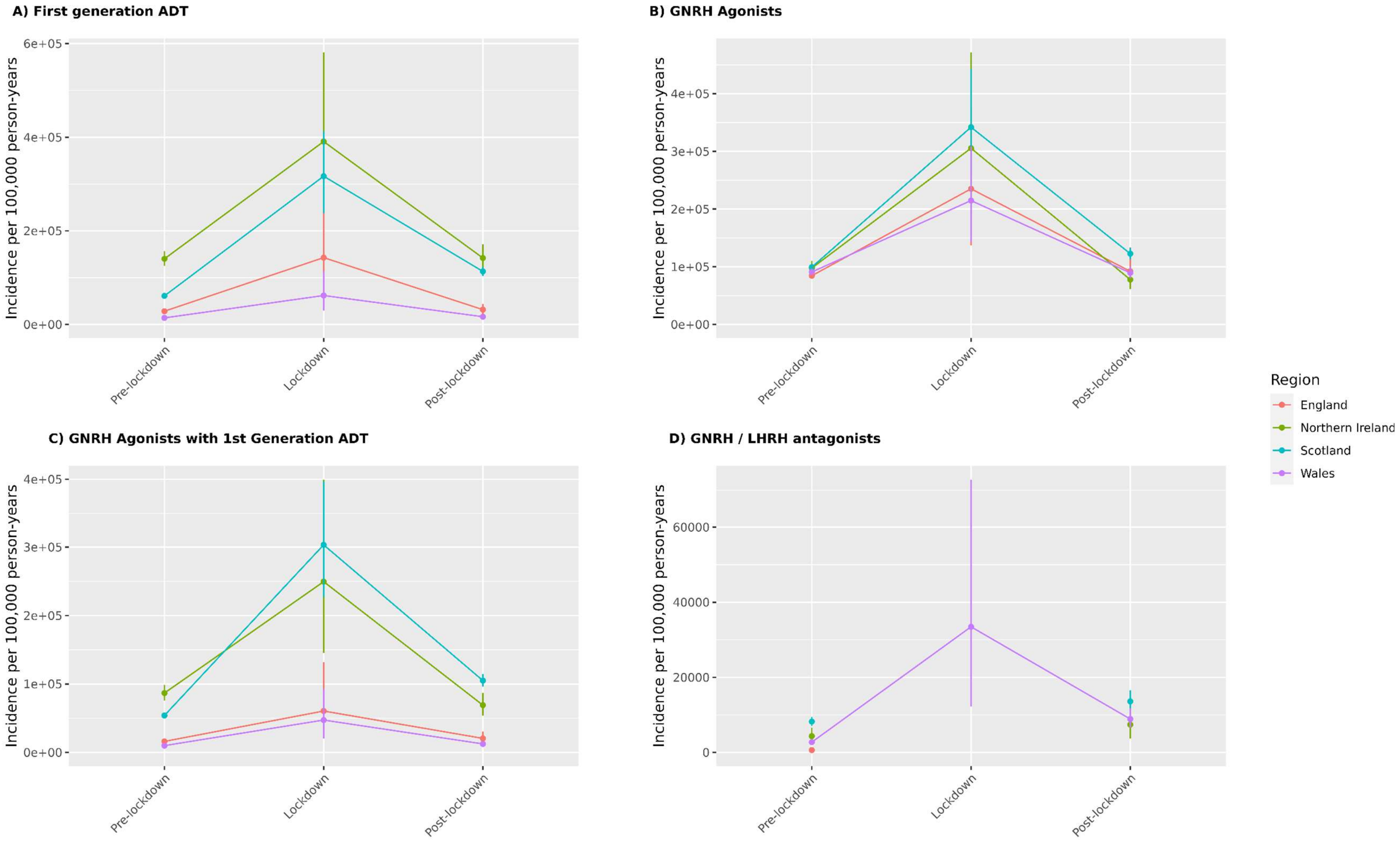


**Note.** ADT = Androgen Deprivation Therapy; GnRH = Gonadotrophin releasing hormone; LHRH = Luteinising hormone-releasing hormone.
